# Can health information and decision aids decrease inequity in health care? A systematic review on the equality of their effectiveness

**DOI:** 10.1101/2024.09.24.24314314

**Authors:** C Ellermann, J Hinneburg, C Wilhelm, FG Rebitschek

## Abstract

**Objectives:** Systematic review of studies evaluating evidence-based health information (EBHI) and decision aids (DAs) in terms of the extent to which inequity-producing factors have been considered and how these factors affect access to health-related information and informed decision-making.

**Study design:** Systematic review of randomised controlled trials.

**Methods:** Systematic searches were performed in the Cochrane Library, PubMed, Embase, PsycINFO, CINAHL, ERIC and PSYNDEX from inception to May 2023 to identify evaluation studies of EBHI and DAs that take into account factors associated with unequal opportunities as defined by PROGRESS Plus. Information on the effect of these factors was extracted and analysed in terms of outcomes relevant to the decision-making process.

**Results:** Few studies have examined the impact of EBHI/DAs on outcomes relevant to decision-making with respect to inequity-producing factors. In our final synthesis,12 studies were included. A positive association between the effectiveness of the intervention and the disadvantage status could be found twice and a negative association in three studies. Overall, most of the studies found no difference in knowledge gain, decision conflict and shared decision-making (SDM) between those advantaged and disadvantaged in terms of ethnicity, gender, education, age, income, health literacy, numeracy or socioeconomic status (SES). However, few trials examined this effect and the effect was considered solely in subgroup analyses that were probably underpowered, so asymmetries between these groups may not have been detected in the existing designs.

**Conclusion:** EBHI and DAs have been shown to be effective in promoting decision-making and thus in improving health care. To improve health care equitably, greater attention needs to be paid to methodological requirements in evaluations to fully capture potential differences in access to health-related information between individuals or in populations within the target groups of EBHI/DAs.

**PROSPERO registration:** CRD42018103456

**What is already known?:**

- There is evidence that EBHI and DAs do not reach certain patient groups because while being developed and evaluated they do not adequately take into account differences in access to health-related information between different social groups.
- There is insufficient evidence whether EBHI and DAs are equally effective for people with factors that are more or less associated with equal access to health information.

**What the study adds:**

- A systematic review of evaluation studies of EBHI and DAs to consider factors that lead to inequity and analysis of how these factors influence the intervention effects in terms of access to health-related information and outcomes relevant for decision-making.

**How this study might affect research, practice or policy?:**

- Our research makes a valuable contribution to more equitable health care by stressing critical inequality factors that may influence informed decision-making with the help of EBHI and DAs.

## Introduction

Evidence-based health information (EBHI) and decision aids (DAs) have the potential to reduce inequity by providing equal access to relevant information. They aim to facilitate informed decision-making by providing all those facing a health decision with sufficient knowledge, with which people can make a decision in line with their treatment preferences ^[1]^. Unlike EBHI, DAs directly aim to elicit preferences and support patient decision-making by providing decision-makers with detailed and personalised options and outcomes ^[2]^. EBHI and DAs are developed in an extensive process that involves continuous adaptation ^[3]^. Information needs and requirements of the target group are crucial throughout the development process ^[4]^. However, the group targeted by the general decision-making situation can be very heterogeneous in terms of their individual ability to understand and process information and to draw conclusions. Accordingly, there is a risk that those individuals or groups within the target group who are already disadvantaged in terms of informed and shared decision-making (SDM) will benefit least from EBHI and DAs. It has long been recognised that patient education materials fail to adequately address differences in access to health-related information between different social groups by not taking sufficient account of factors that are associated with inequalities ^[5–7]^. These factors include race/ethnicity, language, gender, education, socioeconomic status (SES) and age, which, according to PROGRESS-Plus, lead to unequal opportunities and thus differences in health outcomes ^[8]^. Many of the factors influence access to health care in general. For instance, written information is often at a higher reading level or developed for general audiences and therefore does not reach people with low literacy skills, education, and SES or those from diverse cultural groups ^[7, 9, 10]^, although the checklist of quality criteria from the International Patient Decision Aid Standards (IPDAS) has always included the use of plain language ^[11]^.

At present, it is unclear whether EBHI and DAs are equally effective for all patient groups or which subgroups benefit (most), as few trials and systematic reviews have investigated this effect. However, evidence exists that disadvantaged patients (e.g. with lower literacy skills, education and socioeconomic status) are less likely to make informed choices and are more likely to regret their decision than advantaged groups ^[7]^. Furthermore, a patient-level meta-analysis based on seven unpublished RCTs from the Knowledge and Evaluation Research Unit of the Mayo Clinic, USA, suggests that DAs during the clinical encounter lead to a greater increase in risk knowledge in patients with higher education than in those with lower education ^[12]^. Subgroup effects on the basis of race were imprecise. The most current Cochrane Review published in January 2024 ^[2]^, did not investigate whether the positive effect of DAs applies equally to all patient subgroups. This gap is the subject of our review. However, the authors of the review noted that more robust evidence that decision aids can improve health equity or reduce inequalities in access to care could further support their use in clinical practice, which is currently rare ^[2]^.

Our review aims to systematically assess to which extent studies evaluating the effectiveness of EBHI/DAs have considered factors that lead to inequalities in access to health-related information. We also analyse how their effectiveness in terms of decision outcomes varies within the target groups according to these factors.

## Methods

The reporting follows the Preferred Reporting Items for Systematic Reviews and Meta-Analysis (PRISMA) 2020 ^[13]^ and PRISMA-Equity statement ^[14]^ to account for equity-related aspects. PROSPERO has been used for prospective registration (CRD42018103456).

### Study inclusion and exclusion

Studies were included: (1) if they compared the effectiveness of EBHI/DAs (intervention) with usual care or no information (control) between different social groups in a RCT, (2) when a specific health decision needed to be made, and (3) if the main outcome of interest was an informed decision or a related aspect.

Therefore, we also included studies that analysed effects on knowledge, attitude, uptake and related outcomes such as decision concordance, risk perception, recall, understanding, intention, SDM, decision conflict and regret. Furthermore, studies (4) had to include an analysis of the effects based on at least one characteristic that stratifies health opportunities and outcomes according to the PROGRESS-Plus definition (place of residence, race/ethnicity/culture, occupation, sex, religion, education, SES, social capital, age, disability and sexual orientation) in their results section. We also included non-EBHI/DA-interventions, as we expected few studies to examine the effectiveness of EBHI/DAs in different social groups. These studies had to provide information about the benefits and harms of the treatment options (e.g. participation vs. non-participation) for those about to make a decision – one of the minimum requirements for EBHI ^[15]^.

We excluded: (1) studies comparing different formats of EBHI/DAs with the same content (e.g. tabular vs. written information), (2) studies focusing on a concrete disadvantaged group (e.g. people with low literacy), and (3) studies including a multicomponent intervention, where the intervention effect is not clearly attributable to the (non-)EBHI/DA.

### Search strategy, study selection and data extraction

We searched the Cochrane Library, MEDLINE and PubMed (via PubMed), EMBASE (via Ovid), PsycINFO (via Ovid), Cumulative Index to Nursing and Allied Health Literature (CINAHL) (via Ovid), ERIC and PSYNDEX (via Ovid) from inception to May 2023 using a combination of English-language search terms including or describing determinants and factors of health inequality in combination with the intervention and outcome (***Supplement 1***). EndNote 20 was used for reference management.

After duplicates were removed, two reviewers independently reviewed all titles and abstracts via the browser application Rayyan ^[16]^. The first reviewer was always the first review author (CE); the second reviewer varied (CH, JB, JH). Studies included were then screened in full text by two reviewers independently, with disagreements resolved by discussion. Additionally, reference lists of included studies and of relevant systematic reviews identified were checked for grey literature.

A standard data extraction form was used to extract data, focusing on which aspects of inequality, as defined by PROGRESS Plus, were taken into account and the impact of each aspect for the respective factor.

### Risk of Bias

Two people independently assessed the risk of bias using Cochrane’s ROB 2.0 ^[17]^ tool for RCTs. The first person was always the first review author (CE); the second person varied (CM, DA, JB, PB). Disagreements were resolved by discussion and, if necessary, with the involvement of a third person (FGR).

### Data synthesis

Due to the lack of relevant reported data and heterogeneous instruments to access outcomes (e.g. informed choice) and disadvantage level of study participants (e.g. health literacy (HL)) and to the effect of outcomes being available for only some subgroups in the identified studies, the Grading of Recommendations Assessment, Development and Evaluation (GRADE) system ^[18]^ could not be applied and a meta-analysis was not possible. The results were therefore summarised narratively and effects visualised using harvest plots ^[19]^, including only studies with low and medium risk of bias.

### Patient and public involvement

We did not involve patients in the design of the trial or in the dissemination of the results.

## Results

The literature searches identified 3803 studies, of which 216 potentially relevant studies were included for full-text screening. Data extraction was performed for seven studies. Five additional studies were identified from the reference lists of the studies included and of relevant systematic reviews. The study selection process is shown in ***Figure 1***. Sixty studies that were initially considered eligible were excluded due to wrong intervention (e.g. multicomponent analysis without separate analysis of the EBHI/DAs), 118 studies lacked an equity analysis, eight studies used the wrong study design and ten studies reported on the wrong outcome or population. See the appendix for a list of excluded studies and specific reasons (***Supplement 1***).

**Figure 1:**
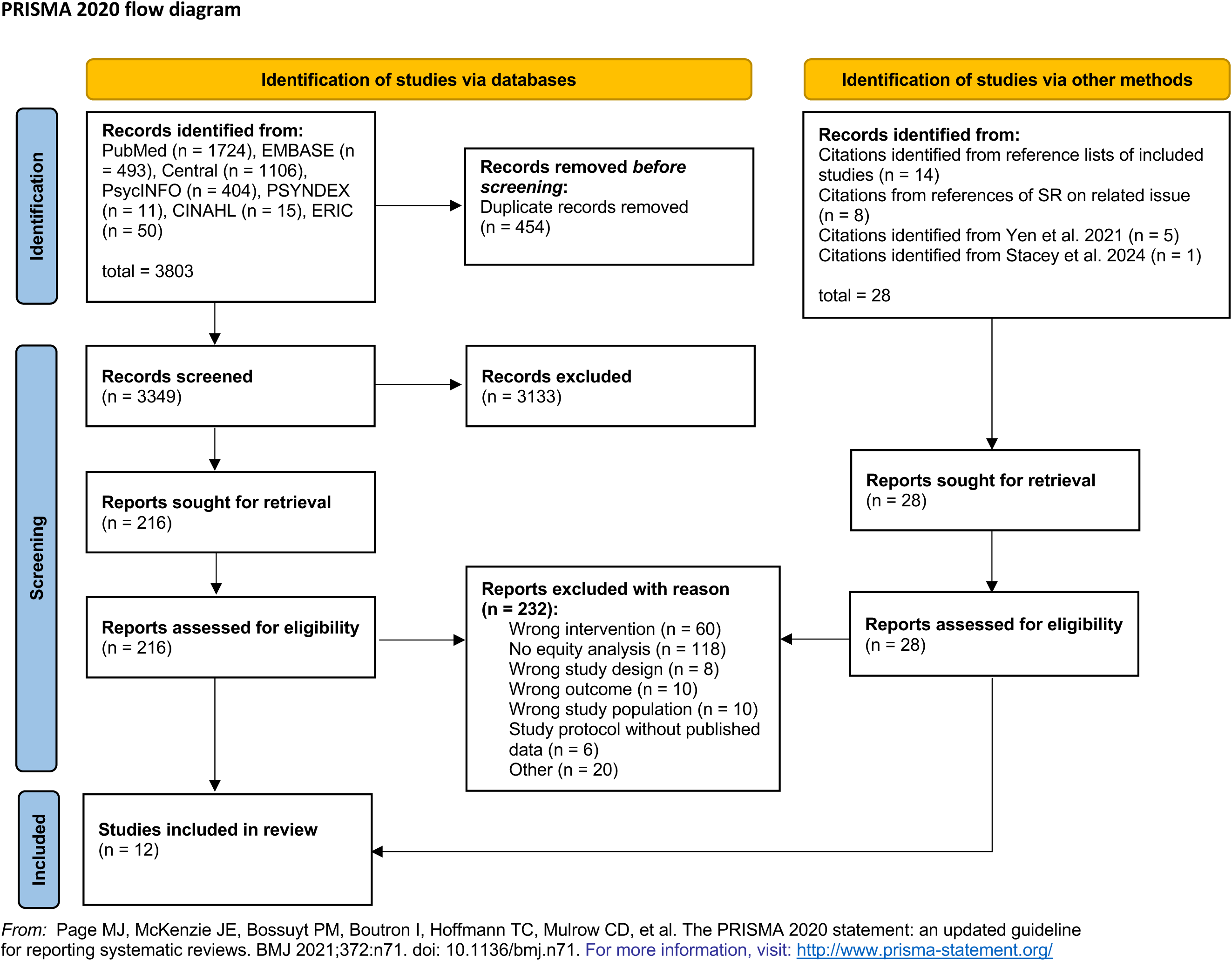
Prisma Flow Chart on study inclusion.

### Characteristics of included studies

Of the 12 trials finally included, nine studies ^[6, 7, 20–26]^ used an intervention developed according to the IPDAS criteria ^[4]^, so were EBHI/DAs; the remaining three ^[27–29]^ were non-EBHI/DAs. Nine studies were carried out in the USA and one study each in the UK, Australia and Canada in heterogenous clinical settings (e.g. specialised medical centres and clinical sites, primary care settings/practices). Two studies showed a thematic overlap in the decision-making situation (prenatal testing) ^[20, 29]^; all other topics occurred only once. Due to the heterogeneous decision-making situations, the target groups of the intervention were also very diverse and ranged from pregnant women to elderly patients with increased risk or pre-existing conditions (e.g. kidney disease, early stage breast cancer), among others. Trial sizes ranged from 60 to 900 participants, meaning that individual subgroups by inequity producing factor were sometimes very small. Further key characteristics of the included publications and trials can be found in ***Table 1***.

**Table 1:** Characteristics of included studies.

| Author/<br>Year | Study<br>design | Country | Setting | Population | Inequity-factor<br>(by PROGRESS<br>Plus) | N | Intervention | Control | Intervention<br>effect<br>assessment | Outcomes of<br>relevance | Risk of<br>Bias |
| --- | --- | --- | --- | --- | --- | --- | --- | --- | --- | --- | --- |
| Durand<br>et al.<br>2021<sup>[7]</sup> | RCT | USA | 7 clinics within<br>4 National<br>Cancer<br>Institute-<br>designated<br>cancer centers | Women with biopsy-<br>confirmed diagnosis of<br>early staged breast cancer<br>(stages I-IIIa) | By<br>socioeconomic<br>status (SES),<br>education | 616 | **Evidence-**<br>**based** (pictorial)<br>Option Grid on<br>breast-<br>conserving<br>surgery and<br>mastectomy | Usual care<br>(surgeons<br>provided their<br>standard<br>information<br>about breast<br>cancer) | Subgroup analysis | Knowledge,<br>decision<br>concordance,<br>SDM, decision<br>regret,<br>treatment<br>choice | **Some**<br>**concerns** |
| Gordon<br>et al.<br>2017<sup>[27]</sup> | RCT | USA | 2 medical<br>centres | Kidney transplant<br>candidates | By age,<br>race/ethnicity,<br>gender,<br>education,<br>income,<br>employment<br>status,<br>HL | 288 | **Non-evidence**<br>**based** iPad app,<br>"Inform Me:<br>about Increased<br>Risk Donor<br>Kidneys"<br>(n=133) | **Control group:**<br>routine<br>transplant<br>education and<br>31-item multiple-<br>choice posttest<br>on paper after<br>(n=155) | Subgroup analysis | Knowledge | **Some**<br>**concerns** |
| Healton<br>et al.<br>1999<sup>[28]</sup> | RCT | USA | Community<br>family<br>planning<br>clinics and<br>hospital-based<br>HIV centres in<br>19 sites from<br>nine cities | Women including Black,<br>Puerto Rican & Non<br>Puerto Rican Latinas | By age,<br>education,<br>race/ethnicity,<br>language<br>preference | 653 | **Non-evidence-**<br>**based** patient<br>brochure on<br>zidovudine<br>therapy during<br>pregnancy to<br>reduce perinatal<br>HIV<br>transmission | No brochure | Subgroup analysis | Knowledge,<br>intention,<br>attitude | **High** |
| Hewison<br>et al.<br>2001<sup>[29]</sup> | RCT | UK | One medical<br>centre and<br>outpatient | Consecutive women<br>referred for antenatal care<br>to the Hull Maternity<br>Hospital | By age | 2000 | **Non-evidence**<br>**based** video<br>information at<br>home for Down<br>syndrome<br>screening<br>(n=993) | **Control:**<br>women who did<br>not receive the<br>screening video<br>at home<br>(n=1007) | Subgroup analysis | Knowledge | **Some**<br>**concerns** |
| Hunter<br>et al.<br>2005<sup>[20]</sup> | RCT | Canada | One children's<br>hospital | Pregnant women aged 35<br>higher at the time of<br>delivery and their partners;<br>gestational age of 18<br>weeks or less | By gender | 352 | **Non-evidence**<br>**based**<br>audiotape-<br>booklet decision<br>aid + option of<br>genetic<br>counselling<br>(n=116) | **Control arms:**<br>1. individual<br>genetic<br>counselling<br>(n=126)<br>2. genetic<br>counselling in a<br>group (n=110) | Subgroup analysis | Knowledge,<br>decisional<br>conflict | **Some**<br>**concerns** |
| Patzer<br>et al.<br>2018<sup>[21]</sup> | RCT | USA | Three<br>transplant<br>centres | Patients living with end-<br>stage renal disease<br>(ESRD); 18 to 70 years of<br>age, no previous solid | By ethnicity,<br>gender,<br>HL,<br>numeracy | 443 | **Evidence-**<br>**based** web-<br>based DA about<br>kidney | Centre-specific<br>standard of care;<br>education about<br>kidney | Subgroups<br>analysis | Knowledge | **High** |
|  |  |  |  | organ or multiorgan transplant |  |  | transplant compared to dialysis | transplant only |  |  |  |
| Rising et al. 2018<sup>[6]</sup> | Multicentre RCT | USA | 6 Emergency departments | Patients included adults (17 years of age) who presented to the ED with a chief complaint of chest pain, had an initial negative cardiac workup | By age, gender, ethnicity, education, numeracy, HL, Income, health insurance | 898 | **Evidence-based** Chest Pain Choice (COC) decision aid | Usual care | Subgroup analysis (secondary analysis) | Knowledge, decisional conflict, SDM | **Low** |
| Singh et al. 2019<sup>[22]</sup> | Multicentre RCT | USA | Clinical settings (in- and outpatients) | Women with lupus nephritis | By race/ethnicity/ language, education, income, numeracy, graphical literacy, HL | 298 | **Evidence-based** decision aid regarding lupus nephritis and its treatments | Paper pamphlet on lupus kidney disease from the American College of Rheumatology | Subgroup analysis | Decisional conflict, informed choice | **Some concerns** |
| Skains et al. 2019<sup>[30]</sup> | RCT | USA | Seven clinical sites | Parents of children with minor head trauma | By education, HL, numeracy, household income, SES | 971 | **Evidence-based** decision aid for parents of children with minor head trauma (n=493) | Clinicians proceeded with their usual SDM discussion with parents (n=478) | Subgroup analysis (secondary analysis) | Knowledge, decisional conflict, SDM | **Some concerns** |
| Thomas et al. 2013<sup>[24]</sup> | RCT | USA | 3 medical centres | Patients eligible for a primary prevention implantable cardioverter/defibrillator (ICD) | By race/ethnicity | 59 | **Evidence-based** educational video on ICD | Health care provider counselling (usual care) | Subgroup analysis | Knowledge, decisional conflict, uptake/ intention (decision-making) | **High** |
| Trevena et al. 2008<sup>[25]</sup> | RCT | Australia | Six primary care locations | People aged 50-74 years deciding whether to undergo colorectal cancer screening | By education | 314 | **Evidence-based** decision aid for colorectal cancer screening | The consumer version of Australian guidelines | Subgroup analysis | Knowledge, informed choice | **Some concerns** |
| Williams et al. 2012<sup>[26]</sup> | RCT | USA | Georgetown University Medical Center and Howard University Cancer Center | Men pre-registered for PSA screening with no history of prostate cancer | By race/ethnicity |  | **Evidence-based** decision aid on PSA screening delivered at home or at the clinic | Usual care delivered at home or at the clinic | Subgroup analysis | Knowledge, decisional conflict | **High** |

### Risk of bias of included studies

Risk of bias was assessed for the individual trials at the study level. Only two out of the 12 trials were judged to be at low risk of bias across all domains and three of the trials as having a high risk of bias in at least one area (***Figure 2*)**. Five studies each were classified as having some concerns with regard to the randomisation process or the selective reporting of study results. For instance, although most trials were registered, information on a priori planned analyses and other details were often missing.

**Figure 2:**
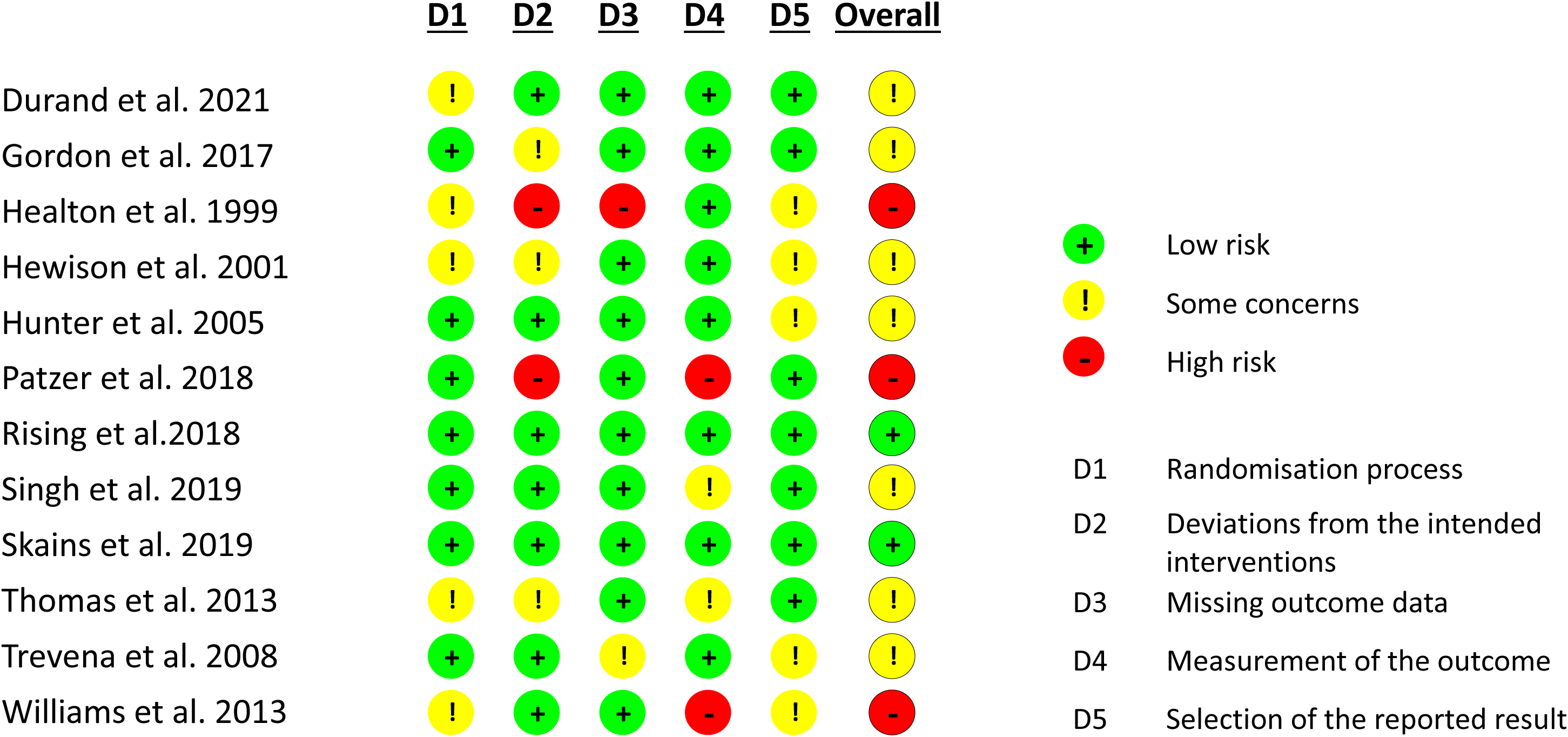
Risk of bias of included studies (N=12).

Crucially, none of the 12 trials was initially designed to assess the effect of the intervention on different social groups; most of the trials assessed the effect through subgroup analyses by social group. This often led to very small numbers of participants for the subgroups, suggesting that the trials were insufficiently powered to detect an intervention effect. Therefore, the overall quality of the evidence is rated as moderate.

### Outcomes relevant for assessing the intervention effect

Most frequent outcomes were knowledge in 11 out of the 12 studies and decisional conflict in six studies (***Supplement 2 - table S1***). Intention/uptake, SDM and informed decision/decision concordance were outcomes of interest in three studies each, decision regret and attitude were outcomes in one study each.

### Consideration of inequity-creating factors

Five inequity-producing factors (as according to PROGRESS-Plus) were considered at least once (***Table 2***). Six out of 12 studies considered more than one factor but only two ^[7, 30]^ considered intersectionality, which refers to the overlapping of various disadvantages (e.g. low-level education and cultural background) ^[31]^. In one of the studies, the authors defined study participants as socioeconomically disadvantaged if they were non-white, had low HL or numeracy skills and had a low income (< USD 40,000) ^[30]^. In the second study ^[7]^, patients with lower SES were defined as having at least two of the following characteristics: lower income, lower education or underinsurance.

**Table 2:**
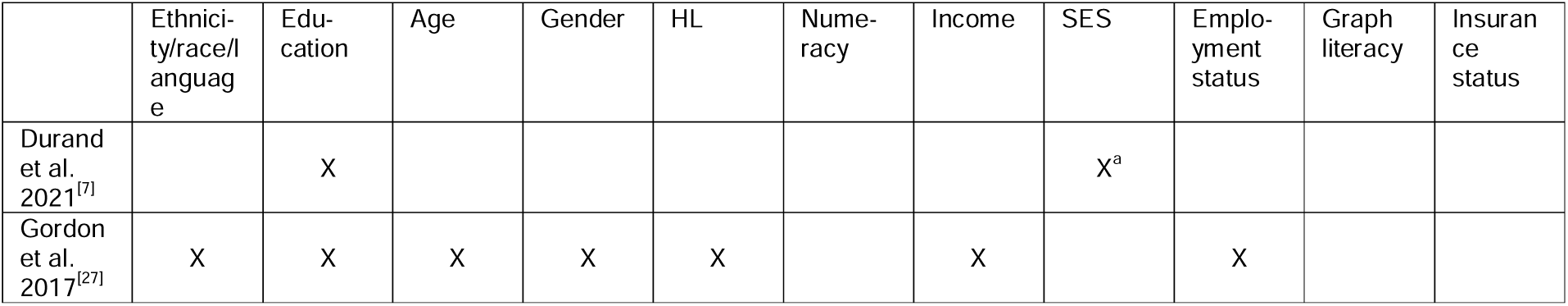
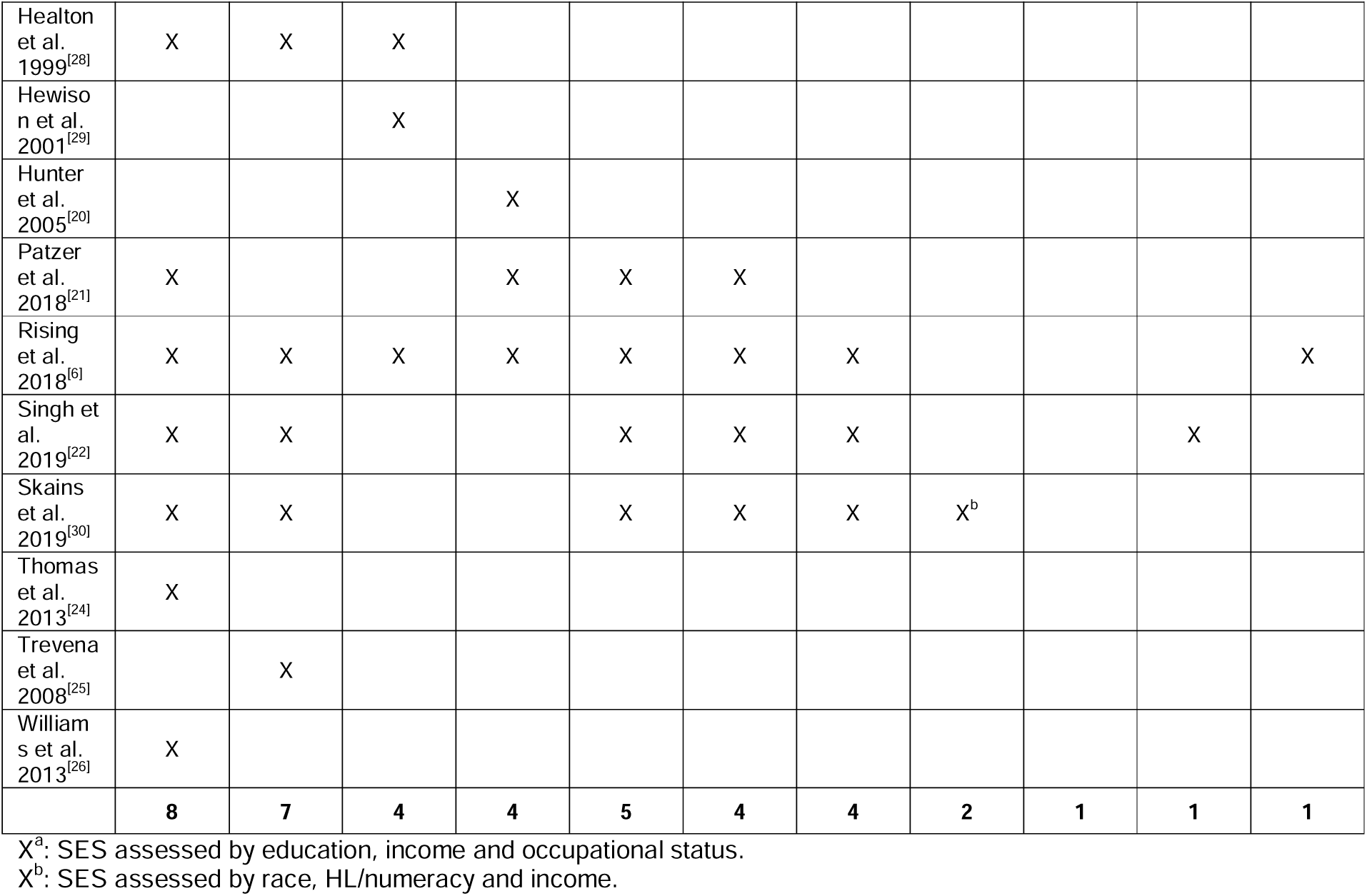
Sociodemographic factors used in the included studies (N = 12) to assess the intervention effect in subgroups.

Only one further study ^[28]^ mentioned that one of the subgroups (non-Puerto Rican Latinas) might benefit the least, as they had the lowest level of English skills and some did not even speak Spanish.

The intervention effect was most frequently observed according to ethnicity/language preference (***Table 2***). The most commonly assessed outcomes by inequality factors were knowledge, decisional conflict and SDM (***Supplement 2 - Table S2***). The results for the inequality factors are only summarised below for these three factors, as the number of studies for other factors is very small, making it almost impossible to draw conclusions.

### Synthesis of intervention effect by inequality factors

Overall, among heterogeneous studies in terms of decision-relevant outcomes and inequality factors only three studies mentioned group-specific intervention effectiveness ^[6, 7, 22]^. The results are summarised narratively for each inequality factor. Only studies with low and medium risk of bias are presented in the forest plots for the most frequent endpoints (***Figure 3 A-C***).

**Figure 3 A-C:**
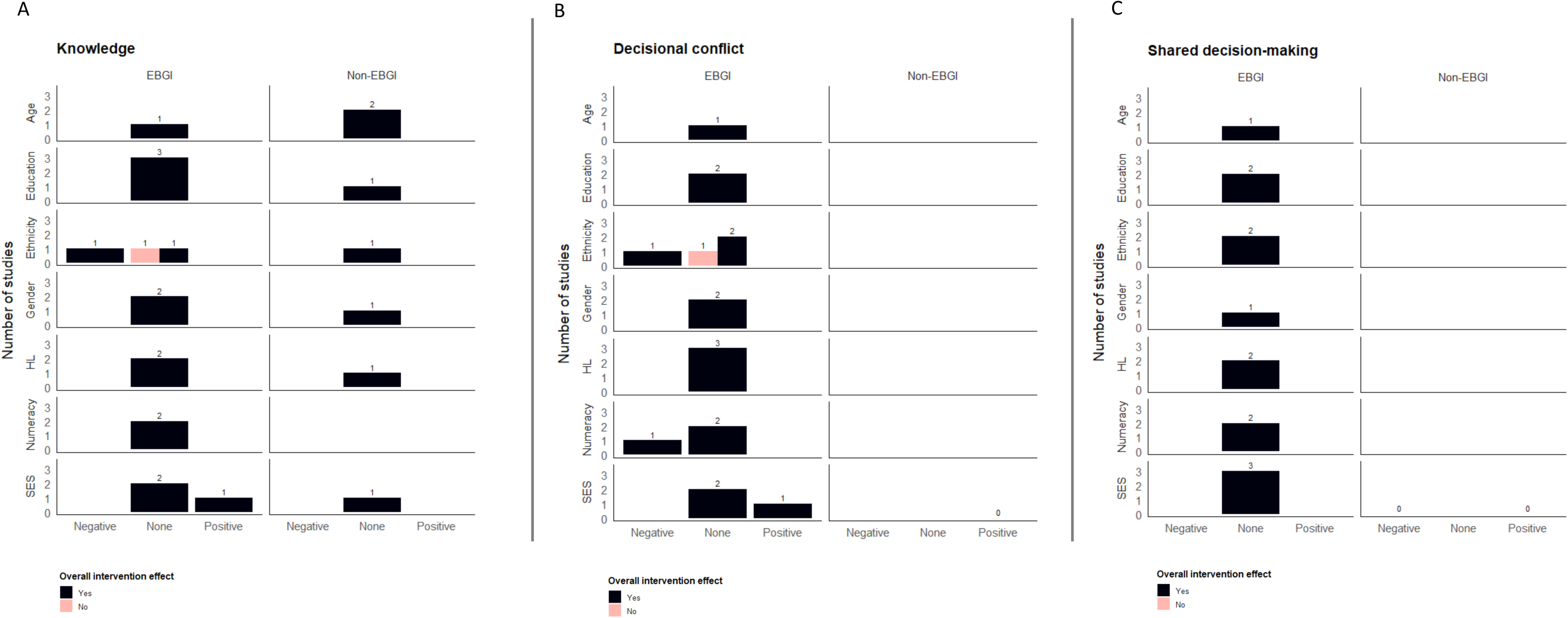
Harvest plots showing the number of intervention studies that either negatively, positively, or neither affect knowledge change, decisional conflict or SDM by inequality factors. Overall intervention effect “No” means that an intervention was not shown to be effective for the full sample with regard to knowledge, decisional conflict or SDM. Studies with high risk of bias were excluded.

#### Age

Three trials ^[6, 28, 29]^ tested for a group-specific intervention effect that was indicated in one high-risk-of-bias study alone (only one knowledge aspect) ^[28]^. Another trial reported higher knowledge scores after intervention across four different age groups without indication of differential effectiveness ^[27]^. Age groups do not seem to be disadvantaged by interventions, whether EBHI/DA or not (***Figure 3 A-C***), but the lack of pre-post assessments across intervention groups and heterogeneous categorisations of age groups may have concealed such differences.

#### Education

None of the four trials that analysed knowledge effects confirmed an association with education ^[6, 25, 28, 30]^ (***Supplement 2 – table S4***). A fifth revealed similar knowledge gains ^[27]^ (***Figure 3 A-C***). Any EBHI/DA seemed to be equally effective for people with higher and lower levels of education for SDM ^[6, 30]^, decisional conflict ^[22, 30]^, attitudes ^[28]^, intention ^[28]^ and informed choice ^[22]^, although each of these endpoints was assessed in a maximum of two studies. A single study ^[7]^ found that the intervention increased disparities in decision regret between those with and without high school diploma.

#### Ethnicity/race and language preference

Among trials where ethnicity was taken into account (***Supplement table S5***), one ^[6]^ indicated that the intervention was more effective for white patients than for other ethnic groups (e.g. African Americans, Latinas or other racial/ethnic groups) in terms of knowledge (***Figure 3 A***). The study, however, lacked an adjustment for multiple testing. Three high-risk-of-bias trials had contradictory results regarding differential effectiveness ^[21, 26, 28]^. Two further trials ^[27, 30]^ found similar knowledge gains across ethnicities, even though Gordon et al. ^[27]^ suggest – without a statistical test – that knowledge improved more in non-Hispanic whites than in other non-African American racial/ethnic groups. A seventh trial did not show any knowledge effect ^[24]^. Interventions were equally effective for white and other ethnic groups with regard to decisional conflict and SDM ^[6, 30]^ (***Figure 3 B-C***). A study with high risk of bias also suggested equal effects with regard to behavioural intention ^[28]^. Singh et al. ^[22]^, however, suggest advantages for whites in the effects on informed choice and decisional conflict, thereby indicating a negative association (***Figure 3 B***). Although this finding for language preference was confirmed in the same study, participants preferring Spanish, who had a presumably high overlap with the Hispanic groups, were less represented in the sample, which might indicate a power problem.

A high-risk-of-bias trial that looked at the effect of the intervention by language preference also indicated that those who received the intervention in their preferred language/mother tongue (English-speaking or bilingual participants) knew more than those with some language barriers (Spanish-speaking participants) ^[28]^.

#### Gender

No difference between women and men regarding intervention effects on knowledge, decisional conflict and SDM was indicated by any study ^[6, 20, 21, 27]^ (***Figure 3 A-C; Supplement table S6***). Two studies ^[20, 21]^, one ^[21]^ with high risk of bias, assessed baseline decisional conflict and knowledge. Only one study ^[6]^ tested the respective interactions with gender.

#### Health literacy

One study showed higher decision-relevant knowledge with intervention for those with adequate, moderate and inadequate literacy skills ^[27]^, but neglected to report the interaction between these groups (***Supplement 2 – table S7***). Two studies ^[6, 30]^ that observed the interaction between having a low or typical HL with the intervention could not reveal differences in knowledge (***Figure 3 A***). However, the statistical power does not seem to be sufficient to detect such an interaction if it exists. In one trial with high risk of bias, people with high literacy skills appeared to achieve greater knowledge scores than did those with lower literacy ^[21]^. Each intervention was equally effective for SDM ^[6, 23]^ and decisional conflict ^[6, 22, 30]^ in studies of people with low and typical HL (***Figure 3 B-C***).

#### Numeracy

Knowledge was suggested to increase more for those with medium or high numeracy scores than for those with lower scores in one trial with high risk of bias ^[21]^ (***Supplement 2 – table S7***). Two further studies ^[6, 30]^ do not support this unambiguously in their comparisons of groups with low vs. typical numeracy (***Figure 3 A***). In one study ^[6]^, there was no adjustment for multiple testing despite 80 comparisons, while both studies may not have been adequately powered to test for the interaction of numeracy with the intervention effects.

For decisional conflicts, one trial ^[22]^ showed that, compared to usual care, the intervention was effective only for people with higher numeracy skills, which indicates a disparity increase (***Figure 3 B***). No difference in terms of both decisional conflict and SDM could be observed in the two further trials considering numeracy ^[6, 23]^ (***Figure 3 B-C***).

#### SES

Intervention effects by socioeconomic factors have been investigated with the help of the SES (two studies ^[7, 30]^), income (four studies ^[6, 22, 27, 30]^) and employment status (one study ^[27]^) (***Supplement 2 - table S8***). Durand et al. ^[7]^ found that the difference in decision-relevant knowledge after an effective intervention between those with lower and higher SES was smaller than in the control group (***Figure 3 A***). The other study could not confirm that finding ^[30]^. Nor did four further studies, indicating similar knowledge gains across a broad income spectrum ^[6, 27, 30]^ (below 25k, below 65k, above 65k US dollars per year) and independent of employment status ^[27]^. None of the studies ^[6, 7, 22, 30]^ found an effect on decisional conflict and SDM. Furthermore, no difference was found between people with lower and higher SES ^[7, 30]^ in terms of intervention effects on decision concordance, decisional conflict, SDM and treatment choice. Nor could an interaction effect of income with interventions on decisional conflict and SDM be confirmed ^[6, 30]^. Only Singh et al. ^[22]^ found largest effectiveness on decisional conflict on those who reported lowest annual income (<40k). Across all studies we found, this is the only evidence that an EBHI/DA-intervention – and, more generally, any health information intervention – can reduce disparities in improving decisional conflict (***Figure 3 B***). Overall, only economic indicators of inequality revealed positively associated intervention effects.

## Discussion

### Principal findings

Our systematic review reveals that inequity-producing factors are rarely considered in effectiveness studies of EBHI/DAs. Included trials took into account at least one factor as according to PROGRESS-Plus, but looked at the effect only in subgroup analyses. The designs were heterogeneous, with few directly testing the association between intervention effects and the disadvantage status, and few assessing baseline levels of the respective outcomes. Sample sizes were often small, and for many of the subgroups, few trials could be identified that provided evidence to draw conclusions about effectiveness and/or the data were poorly reported.

In terms of knowledge, not everyone benefits equally, which is a key endpoint for informed decision-making and SDM. People who were considered disadvantaged because of their ethnicity (e.g. African Americans, Latinas, or other racial/ethnic groups) or language in at least one study attained less knowledge in most studies, which may have prevented the identification of asymmetries in both knowledge acquisition and other outcomes. For decisional conflict, ethnic groups seem to have benefitted equally in three out of four studies, lower and higher health numerates did so in three identified studies, men and women in two identified studies, lower and higher educated in two and younger and older age groups in one study. One of three identified studies analysing numeracy and one out four studies analysing ethnicity respectively showed a disadvantage for low numerates and non-whites in terms of decisional conflict by the intervention. One of three trials analysing SES showed that knowledge inequalities between people of different SES can be reduced by providing EBHI/DAs. No differences in the effect for SDM by ethnicity, education, gender, income, HL or numeracy could be identified across all three studies. Again, however, because only few studies were available and this effect was only considered in subgroup analyses with presumably inadequate power, asymmetries across inequality factors may have not been recognised.

### Strengths and limitations

Through our research with the best available evidence, it has become clear that inequity-producing factors were rarely considered in planning the evaluation and that methodological approaches were not sufficient to make statements about the effect between and within different groups.

Potential limitations: First, despite our efforts to search extensively for RCTs of EBHI/DAs that evaluated the effectiveness in different social groups, our search possibly missed relevant RCTs that met our inclusion criteria. Studies may not have been identified in the title-abstract screening, because the effect of inequity-producing factors was often only analysed in subgroups or mentioned in the discussion only.

Our focus on RCTs may have neglected observational evidence that is more inclusive of disadvantaged subgroups. Because most studies made no assumptions about possible differences in effectiveness due to their retrospective analysis of social factors, we did not define a priori a classification of disadvantaged or not disadvantaged.

Having also included trials without information prepared according to quality criteria for DAs ^[4]^ and not specifying a minimum requirement in the study protocol for those studies, we subsequently specified that trials with non-EBHI had to report at least the benefits and harms. However, we did not assess the quality of the individual DAs and instead relied on the statement that the DAs had been developed according to certain criteria. Thus, some of the DAs may not meet the highest quality standards of EBHI/DAs.

### Comparison with other studies

Unlike previous systematic reviews, ours focused on systematically assessing and analysing the effectiveness of EBHI and DAs between and within patient subgroups. A systematic review from the Cochrane Review versions on the effectiveness of DAs that included studies until 2021 ^[2]^ found that in only 12% of the included RCTs were the needs of people with low HL or other disadvantaged groups considered in the design and testing of DAs ^[11]^, thereby suggesting differences in the degree of effectiveness between and within patient subgroups. However, the review did not investigate this. A systematic review from 2016 ^[32]^ found that just 17 out of 39 included RCTs in the US included disadvantaged people while developing DAs on cancer screening and treatment. Only 14 of the included studies investigated the effect of DAs in disadvantaged groups (mainly by education and HL), most of them for one specific group and few for and between different social groups ^[32]^.

Furthermore, two systematic reviews found that SDM interventions improve for example knowledge, informed choice and SDM for disadvantaged patients (e.g. lower literacy, education and socioeconomic status) – particularly when tailored to the needs of disadvantaged groups ^[5, 33]^. But the authors note the lack of evidence on the effectiveness of decision-making interventions for disadvantaged people compared with non-disadvantaged people, which was our focus. Other systematic reviews examined DA evaluation studies with a particular focus on a group considered disadvantaged (e.g. people with low literacy ^[34]^, older people ^[35]^ or people with racial or ethnic minority background ^[36, 37]^) or focused on disadvantaged populations, but for SDM interventions in general or multicomponent interventions ^[6, 33, 37]^.

#### Implications and future research

Only two studies found positive associations and three studies found negative associations between intervention effectiveness and disadvantaged group status in terms of SES and ethnicity/numeracy. On the one hand, overproportional decision-making benefits in those with lower SES may indicate a kind of “catch-up effect” and a promise of enabling access to evidence-based health communication for those groups. Disproportionately low benefits of those with low numeracy or an underrepresented ethnic background, on the other hand, may reflect disregard of health communication development standards for addressing difficulties with numbers and language. Nevertheless, advantaged and disadvantaged patients appear to benefit equally from EBHI/DAs. Given the small number of trials and the fact that the effect was considered solely in subgroup analyses, which are likely to be underpowered, asymmetries between factors of inequality may not have been detected by the existing designs. Future research should therefore take better account of critical inequality factors when evaluating EBHI and DAs to contribute to more equitable health care.

## Conclusion

Due to the small number of studies and their methodological and qualitative limitations, there is limited evidence on whether EBHI and DA contribute to good healthcare for all patients or whether they benefit only certain subgroups of patients. More attention should thus be paid to the methodological requirements to fully capture potential effects of the diversity of the target groups. This is the only way to prove and reduce inequalities in informed decision-making and ultimately ensure appropriate and equitable health care.

## Contributions

CE, JH and FGR conceptualised and designed the systematic review. CE, JH, CW and FGR screened the studies and extracted the data for different updates of the systematic review. CE and FGR analysed the results. CE drafted the manuscript and made the revisions. CW, JH and FGR critically revised the manuscript. All authors finally approved the manuscript. CE is the guarantor for this review.

We acknowledge Carolin Harder (CH), Chiara Müller (CM), Donja Ameri (DA), Pauline Brunner (PB) and Julia Beckhaus (JB) for their research support (study screening, data extraction and risk of bias assessment).

## Data availability statement

Most of the data and analyses are available in supplementary files. Additional data are available on request from the corresponding author.

## Ethics statements

No ethics approval was required for this systematic review. Patient consent for publication was not applicable.

