## Supplement 1: Tables S1-2 for "Can health information and decision aids decrease inequity in health care? A systematic review on the equality of their effectiveness"

**Search strategy**

| **Cochrane Library** | | | | |
| --- | --- | --- | --- | --- |
| **Block A: Problem/Population** | **Block B:**  **Intervention** | | **Block C:**  **Outcome** | |
| Social and health inequality/inequity | Health information, decision aids, Interventions to improve SDM or informed choice/decision | | Primary: informed choice  Secondary: | |
| #1 MeSH descriptor: [Healthcare Disparities] explode all trees  #2 MeSH descriptor: [Vulnerable Populations] explode all trees  #3 MeSH descriptor: [Minority Groups] explode all trees  #4 MeSH descriptor: [Minority Health] explode all trees  #5 MeSH descriptor: [Health Literacy] explode all trees  #6 MeSH descriptor: [Education] this term only  #7 MeSH descriptor: [Educational Status] explode all trees  #8 MeSH descriptor: [Ethnic Groups] this term only  #9 MeSH descriptor: [Health Equity] explode all trees  #10 (“vulnerable groups” OR “minority groups” OR “minority population” OR “health literate” OR “health literacy” OR “health equity” OR “health inequity” OR “social equity” OR “social inequity”)  #11 #1 or #2 or #3 or #4 or #5 or #6 or #7 or #8 or #9 or #10 | #12 MeSH descriptor: [Consumer Health Information] explode all trees  #13 MeSH descriptor: [Patient Education Handout] explode all trees  #14 MeSH descriptor: [Decision Support Systems, Clinical] explode all trees  #15 MeSH descriptor: [Decision Support Techniques] explode all trees  #16 MeSH descriptor: [Decision Making] explode all trees  #17 ("health information" or "patient information" or "patient leaflet" or “decision aid”):ti,ab,kw (Word variations have been searched)  #18 (((patient or health) and (pamphlet or brochure or flyer or booklet))):ti,ab,kw (Word variations have been searched)  #19 ((decision) and (intervention or tool or video or technique or technology or technologies or app or instrument or program or material)):ti,ab,kw (Word variations have been searched)  #20 #12 or #13 or #14 or #15 or #16 or #17 or #18 or #19 | | #21 MeSH descriptor: [Patient Medication Knowledge] explode all trees  #22 MeSH descriptor: [Attitude] explode all trees  #23 MeSH descriptor: [Attitude to Health] explode all trees  #24 MeSH descriptor: [Perception] this term only  #25 MeSH descriptor: [Comprehension] explode all trees  #26 MeSH descriptor: [Awareness] explode all trees  #27 MeSH descriptor: [Patient Participation] explode all trees  #28 MeSH descriptor: [Decision Making] explode all trees  #29 MeSH descriptor: [Behavior] explode all trees  #30 (knowledge, attitude, perception, comprehension, decision, “informed choice”, “informed decision”, “decisional conflict”, behavior):ti,ab,kw  #31 #21 or #22 or #23 or #24 or #25 or #26 or #27 or #28 or #29 or #30 | |
| Filter:   - Cochrane Reviews, Cochrane Protocols, Trials - publication date Between May 2021 and June 2023 | | | | |
| 18.05.2023 | | | | |
| **PubMed** | | | | |
| **Block A: Problem/Population** | **Block B:**  **Intervention** | | **Block C:**  **Outcome** | |
| Social and health inequality/inequity | Health information, decision aids, Interventions to improve SDM or informed choice/decision | | Primary: informed choice  Secondary: | |
| "Healthcare Disparities"[Majr] OR "Vulnerable Populations"[Majr] OR "Minority Groups"[Majr] OR "Minority Health"[Majr] OR "Health Literacy"[Majr] OR "Education"[Majr] OR "Educational Status"[Majr] OR "Ethnic Groups"[Majr] OR “health inequity”[TIAB] OR “health inequities”[TIAB] OR "health inequality"[TIAB] OR "health inequalities"[TIAB] OR “minorities”[TIAB] OR “social groups”[TIAB] OR “social disparity”[TIAB] OR “social disparities”[TIAB] OR ((vulnerable [TIAB] OR advantaged [TIAB] OR disadvantaged [TIAB] OR underserved [TIAB]) AND (groups [TIAB] OR people [TIAB] OR population* [TIAB] OR patients [TIAB])) | "Consumer Health Information"[Majr] OR "Patient Education Handout" [Publication Type] OR “health information” [TIAB] OR “patient information”[TIAB] OR “patient leaflet” [TIAB] OR "Decision Support Systems, Clinical"[Majr] OR "Decision Support Techniques"[Mesh] OR "Decision Making"[Mesh] OR ((patient [TIAB] OR health [TIAB]) AND (pamphlet [TIAB] OR brochure [TIAB] OR flyer [TIAB] OR booklet [TIAB])) OR “decision aid”[TIAB] OR “facts box”[TIAB] OR ((decision*[TIAB]) AND (intervention*[TIAB] OR tool [TIAB] OR video [TIAB] OR technique*[TIAB] OR technology [TIAB] OR technologies [TIAB] OR app [TIAB] OR instrument* [TIAB] OR program [TIAB] OR material*[TIAB])) | | "Patient Medication Knowledge"[Majr] OR "Knowledge"[Mesh] OR "Health Knowledge, Attitudes, Practice"[Majr] OR "Perception"[Mesh] OR "Comprehension"[Mesh] OR "Patient Participation"[Mesh] OR "Decision Making"[Mesh] OR “risk literacy”[TIAB] OR attitude [TIAB] OR awareness [TIAB] OR perception [TIAB] OR knowledge [TIAB] OR “patient participation” [TIAB] OR “patient involvement” [TIAB] OR  “shared-decision” [TIAB] OR “decisional conflict” [TIAB] OR “decision regret” [TIAB] OR behaviour [TIAB] OR behavior [TIAB] OR “informed decision”[TIAB] OR “informed decisions” [TIAB] OR “informed choice” [TIAB] OR “informed choices” [TIAB] OR “decision-making”[TIAB] | |
| Filter: RCT  19.05.2023 | | | | |
| **PsycINFO (via EBSCO)** | | | | |
| **Block A: Problem/Population** | **Block B:**  **Intervention** | | **Block C:**  **Outcome** | |
| Social and health inequality/inequity | Health information, decision aids, Interventions to improve SDM or informed choice/decision | | Primary: informed choice  Secondary: | |
| S1 (DE "Disadvantaged") OR (DE "At Risk Populations") OR (DE "Minority Groups") OR (DE "Social Equity") OR (DE "Social Equality") OR (DE "Social Groups") OR (DE "Racial and Ethnic Groups")  OR  S2 TI "Minority Health" OR AB "Minority Health"  OR  S3 TI "health disparit*" OR AB "health disparit*"  OR S4 TI "health inequ*" OR AB "health inequ*"  OR  S5 TI ( (vulnerable OR social OR advantaged OR disadvantaged OR underserved) ) AND TI ( (group or population or people or patient) )  OR  S6 AB ( (vulnerable OR social OR advantaged OR disadvantaged OR underserved) ) AND AB ( (group or population or people or patient) )  OR S7 TI "social disparit*" OR AB "social disparit*"  S8  (S1 OR S2 OR S3 OR S4 OR S5 OR S6 OR S7) | S9 ((DE "Decision Support Systems") OR (DE "Health Information"))  OR  S10 TI ( patient OR health ) AND TI ( information or leaflet or pamphlet or booklet or flyer or handout or brochure )  OR  S11 AB ( patient OR health ) AND AB ( information or leaflet or pamphlet or booklet or flyer or handout or brochure )  OR  S12 TI "fact box" OR AB "fact box"  OR  S13 TI "decision aid" OR AB "decision aid"  S14 S9 OR S10 OR S11 OR S12 OR S13 | | ((DE "Health Knowledge") OR (DE "Risk Perception") OR (DE "Perception") OR (DE "Comprehension") OR (DE "Attitudes") OR (DE "Awareness") OR (DE "Decision Making") OR (DE "Behavior"))  OR  TI "shared decision" OR AB "shared decision"  OR TI "informed choice" OR AB "informed choice"  OR TI "informed decision" OR AB "informed decision"  OR  TI "patient involvement" OR AB "patient involvement"  OR TI ( decision AND (regret OR conflict) ) OR AB ( decision AND (regret OR conflict) ) | |
| Limiter: Publication Date: 20210501-; RCTs: (by Methodology: empirical study, quantitative study, clinical trial)  15.06.2023 | | | | |
| **PSYNDEX (via EBSCO)** | | | | |
| **Block A: Problem/Population** | **Block B:**  **Intervention** | | **Block C:**  **Outcome** | |
| Social and health inequality/inequity | Health information, decision aids, Interventions to improve SDM or informed choice/decision | | Primary: informed choice  Secondary: | |
| S1 (DE "Disadvantaged") OR (DE "At Risk Populations") OR (DE "Minority Groups") OR (DE "Social Equity") OR (DE "Social Equality") OR (DE "Social Groups") OR (DE "Racial and Ethnic Groups")  OR  S2 TI "Minority Health" OR AB "Minority Health"  OR  S3 TI "health disparit*" OR AB "health disparit*"  OR S4 TI "health inequ*" OR AB "health inequ*"  OR  S5 TI ( (vulnerable OR social OR advantaged OR disadvantaged OR underserved) ) AND TI ( (group or population or people or patient) )  OR  S6 AB ( (vulnerable OR social OR advantaged OR disadvantaged OR underserved) ) AND AB ( (group or population or people or patient) )  OR S7 TI "social disparit*" OR AB "social disparit*"  S8  (S1 OR S2 OR S3 OR S4 OR S5 OR S6 OR S7) | S9 ((DE "Decision Support Systems") OR (DE "Health Information"))  OR  S10 TI ( patient OR health ) AND TI ( information or leaflet or pamphlet or booklet or flyer or handout or brochure )  OR  S11 AB ( patient OR health ) AND AB ( information or leaflet or pamphlet or booklet or flyer or handout or brochure )  OR  S12 TI "fact box" OR AB "fact box"  OR  S13 TI "decision aid" OR AB "decision aid"  S14 S9 OR S10 OR S11 OR S12 OR S13 | | ((DE "Health Knowledge") OR (DE "Risk Perception") OR (DE "Perception") OR (DE "Comprehension") OR (DE "Attitudes") OR (DE "Awareness") OR (DE "Decision Making") OR (DE "Behavior"))  OR  TI "shared decision" OR AB "shared decision"  OR TI "informed choice" OR AB "informed choice"  OR TI "informed decision" OR AB "informed decision"  OR  TI "patient involvement" OR AB "patient involvement"  OR TI ( decision AND (regret OR conflict) ) OR AB ( decision AND (regret OR conflict) ) | |
| Limiter: Publication Date: 20210501  25.05.2021 | | | | |
| **Embase (via EBSCO)** | | | | |
| **Block A: Problem/Population** | **Block B:**  **Intervention** | | **Block C:**  **Outcome** | |
| Social and health inequality/inequity | Health information, decision aids, Interventions to improve SDM or informed choice/decision | | Primary: informed choice  Secondary: decision regret, decision conflict | |
| 1. exp health disparity/; 2. exp vulnerable population/; 3. exp minority group/; 4. exp minority health/; 5. exp health literacy/; 6. education/; 7. exp educational status/; 8. ethnic group/; 9. social disparity.ab,ti.; 10. social group.ab,ti.; 11. 1 or 2 or 3 or 4 or 5 or 7 or 8 or 9 or 10 | 12. exp consumer health information/; 13. patient education handout.ab,ti.; 14. medical information.ab,ti.; 15. exp patient information/; 16. patient leaflet.ab,ti.; 17. exp decision support system/; 18. exp decision making/; 19. 12 or 13 or 14 or 15 or 16 or 17 or 18 | | 20. exp knowledge/; 21. exp behavior/; 22. exp perception/; 23. exp comprehension/; 24. exp patient participation/; 25. exp decision making/; 26. exp awareness/; 27. exp shared decision making/; 28. decisional conflict.mp.; 29. decision regret.ab,ti.; 30. informed decision making.mp.; 31. informed choice.mp.; 32. 20 or 21 or 22 or 23 or 24 or 25 or 26 or 27 or 28 or 29 or 30 or 31; 33. 11 and 19 and 32 | |
| Limiter: to yr="2018 - 2021"; RCTS  26.05.2021 (No access via Uni Potsdam for the update in 2023) | | | | |
| **Cinahl** | | | | |
| **Block A: Problem/Population** | **Block B:**  **Intervention** | | **Block C:**  **Outcome** | |
| Social and health inequality/inequity | Health information, decision aids, Interventions to improve SDM or informed choice/decision | | Primary: informed choice  Secondary: decision regret, decision conflict | |
| (MM "Health Status Disparities") OR (MM "Healthcare Disparities") OR (MM "Special Populations") OR (MM "Minority Groups") OR (MM "Health Literacy") OR (MM "Educational Status") OR (MM "Ethnic Groups") OR (TI social group* OR AB social group*) OR (TI health inequ* OR AB health inequ*) OR (TI social disparit* OR AB social disparit*) OR TI ( (vulnerable OR advantaged OR disadvantaged OR underserved) AND (group* OR people OR patient* OR population*) ) OR AB ( (vulnerable OR advantaged OR disadvantaged OR underserved) AND (group* OR people OR patient* OR population*) ) | (MM "Consumer Health Information") OR (MH "Teaching Materials/ED/ES/EV/MT") OR (PT consumer/patient teaching materials) OR (TI 'health information' OR AB 'health information') OR (MM "Decision Support Systems, Clinical") OR (MM "Decision Support Techniques") OR (MM "Decision Support Systems, Management") OR (TI ( (patient OR health) AND (pamphlet* OR brochure* OR flyer OR booklet*) ) OR AB ( (patient OR health) AND (pamphlet* OR brochure* OR flyer OR booklet*) )) OR (TI decision aid OR AB decision aid) OR (TI ‘fact* box*' OR AB 'fact* box*') OR (TI ( decision* AND (intervention* OR tool* OR video* OR technique* OR technolog* OR app* OR instrument* OR program* OR material*) ) OR AB ( decision* AND (intervention* OR tool* OR video* OR technique* OR technolog* OR app* OR instrument* OR program* OR material*) ) ) | | (MM "Knowledge") OR (MH "Health Knowledge") OR (MM "Behavior and Behavior Mechanisms") OR (MM "Perception") OR (MM "Cognition") OR (TI comprehension OR AB comprehension) OR (MM "Consumer Participation") OR (MH "Decision Making, Patient") OR (MH "Decision Making, Family") OR (MH "Decision Making, Computer Assisted") OR (MH "Decision Making") OR (TI shared decision making OR AB shared decision making) OR (MM "Decisional Conflict (NANDA)") OR (MM "Decisional Conflict (Saba CCC)") OR (TI decisional conflict OR AB decisional conflict) OR (TI decision regret OR AB decision regret) OR (TI informed decision OR AB informed decision OR TI informed choice OR AB informed choice) | |
| XX.05.2021 (No access via Uni Potsdam for the update in 2023) | | | | |
| **ERIC (ProQuest)** | | | | |
| **Block A: Problem/Population** | | **Block B:**  **Intervention** | | **Block C:**  **Outcome** |
| Social and health inequality/inequity | | Health information, decision aids, Interventions to improve SDM or informed choice/decision | | Primary: informed choice  Secondary: decision regret, decision conflict, knowledge, comprehension, understanding |
| (((patient OR health) AND (pamphlet OR brochure OR flyer OR booklet OR information)) OR ("decision aid" OR "fact box" OR “decision tool” “decision support”)))  OR (Knowledge OR Understanding OR Behavior OR Perception OR Comprehension OR Participation OR “Health literacy” OR decision “Decision Making" OR “decisional conflict" OR "decision regret" OR "informed decision" OR "informed choice") | | | | |
| Limiter: since 2019 | | | | |
| 21.06.2023 | | | | |

**List of studies excluded with reasons**

|  | **Study** | **Exclusion reason** |
| --- | --- | --- |
| 1. | Abujilban, et al. ^[1]^ | The study did not investigate the equality of the intervention effect between different social groups |
| 2. | Alden, et al. ^[2]^ | The study did not investigate the equality of the intervention effect between different social groups |
| 3. | Allen, et al. ^[3]^ | The study did not investigate the equality of the intervention effect between different social groups |
| 4. | Allen, et al. ^[4]^ | Wrong outcome (subgroup effect only on desire for life-sustaining treatment) |
| 5. | Alsaffar, et al. ^[5]^ | The study did not investigate the equality of the intervention effect between different social groups |
| 6. | AlSagheir, et al. ^[6]^ | The study did not investigate the equality of the intervention effect between different social groups |
| 7. | Aremu, et al. ^[7]^ | Does not inform about different treatment options |
| 8. | Arterburn, et al. ^[8]^ | The study did not investigate the equality of the intervention effect between different social groups |
| 9. | Babapour Mofrad, et al. ^[9]^ | No equity analysis on relevant outcome |
| 10. | Bailey, et al. ^[10]^ | Wrong intervention (multicomponent intervention) |
| 11. | Banegas ^[11]^ | Wrong study design |
| 12. | Barton, et al. ^[12]^ | Wrong intervention (multicomponent intervention) |
| 13. | Baykaner, et al. ^[13]^ | Wrong intervention (multicomponent intervention) |
| 14. | Beach, et al. ^[14]^ | Study protocol (NCT01992926) without final publication of study results |
| 15. | Bernat, et al. ^[15]^ | Wrong intervention (no benefit-harm information) |
| 16. | Beulen, et al. ^[16]^ | The study did not investigate the equality of the intervention effect between different social groups |
| 17. | Bjorklund, et al. ^[17]^ | The study did not investigate the equality of the intervention effect between different social groups |
| 18. | Blalock, et al. ^[18]^ | Wrong intervention (comparison of formats) |
| 19. | Blalock, et al. ^[19]^ | The study did not investigate the equality of the intervention effect between different social groups |
| 20. | Botkin, et al. ^[20]^ | The study did not investigate the equality of the intervention effect between different social groups |
| 21. | Boundouki, et al. ^[21]^ | The study did not investigate the equality of the intervention effect between different social groups |
| 22. | Brenner, et al. ^[22]^ | Conference paper on Brenner, et al. ^[23]^ |
| 23. | Brenner, et al. ^[24]^ | Study protocol on Brenner, et al. ^[23]^ |
| 24. | Brenner, et al. ^[23]^ | Vulnerability of study population unclear |
| 25. | Broome, et al. ^[25]^ | The study did not investigate the equality of the intervention effect between different social groups |
| 26. | Brown, et al. ^[26]^ | The study did not investigate the equality of the intervention effect between different social groups |
| 27. | Bucker, et al. ^[27]^ | The study did not investigate the equality of the intervention effect between different social groups |
| 28. | Buhse, et al. ^[28]^ | Wrong intervention |
| 29. | Buhse, et al. ^[29]^ | The study did not investigate the equality of the intervention effect between different social groups |
| 30. | Cadet, et al. ^[30]^ | Wrong study design (subgroup analysis only on intervention group women) |
| 31. | Carhuapoma, et al. ^[31]^ | Study protocol; original study includes wrong intervention (multicomponent intervention) |
| 32. | Carles, et al. ^[32]^ | The study did not investigate the equality of the intervention effect between different social groups |
| 33. | Carlin, et al. ^[33]^ | The study did not investigate the equality of the intervention effect between different social groups |
| 34. | Carré, et al. ^[34]^ | The study did not investigate the equality of the intervention effect between different social groups |
| 35. | Chan, et al. ^[35]^ | The study did not investigate the equality of the intervention effect between different social groups |
| 36. | Chan, et al. ^[36]^ | No detailed information on intervention and control format |
| 37. | Chavarria, et al. ^[37]^ | Wrong outcome (focus on uptake) |
| 38. | Chen and Yu ^[38]^ | Wrong intervention |
| 39. | Chen, et al. ^[39]^ | Wrong intervention (multicomponent intervention) |
| 40. | Chewning, et al. ^[40]^ | The study did not investigate the equality of the intervention effect between different social groups |
| 41. | Clarke, et al. ^[41]^ | Wrong intervention (no benefit-harm information) |
| 42. | Davison, et al. ^[42]^ | The study did not investigate the equality of the intervention effect between different social groups |
| 43. | Delp and Jones ^[43]^ | Wrong intervention (no benefit-harm information) |
| 44. | DeWalt, et al. ^[44]^ | Multicomponent intervention |
| 45. | Diefenbach, et al. ^[45]^ | Wrong intervention (individual risk information) |
| 46. | Dodin, et al. ^[46]^ | Other language |
| 47. | Doll, et al. ^[47]^ | The study did not investigate the equality of the intervention effect between different social groups |
| 48. | Durand, et al. ^[48]^ | Study protocol publication – final study included |
| 49. | Eckman, et al. ^[49]^ | The study did not investigate the equality of the intervention effect between different social groups |
| 50. | Edwards, et al. ^[50]^ | Wrong intervention |
| 51. | Eiriksdottir, et al. ^[51]^ | Wrong study design (mixed methods without evaluation) |
| 52. | El Miedany, et al. ^[52]^ | The study did not investigate the equality of the intervention effect between different social groups |
| 53. | El Morr, et al. ^[53]^ | No detailed information on intervention and control format |
| 54. | Ellison, et al. ^[54]^ | Wrong intervention |
| 55. | Enzinger, et al. ^[55]^ | The study did not investigate the equality of the intervention effect between different social groups |
| 56. | Eschalier, et al. ^[56]^ | The study did not investigate the equality of the intervention effect between different social groups |
| 57. | Eslami, et al. ^[57]^ | Study protocol publication – final study not found |
| 58. | Fisher, et al. ^[58]^ | The study did not investigate the equality of the intervention effect between different social groups |
| 59. | Fraval, et al. ^[59]^ | Wrong patient population |
| 60. | Freed, et al. ^[60]^ | Wrong outcome |
| 61. | Frosch, et al. ^[61]^ | Wrong study design (sequential quasi- experimental design without randomisation) |
| 62. | Gabel, et al. ^[62]^ | Wrong study population |
| 63. | Gagne, et al. ^[63]^ | The study did not investigate the equality of the intervention effect between different social groups |
| 64. | García, et al. ^[64]^ | The information is not intended to prepare a decision (oocyte donation), but is general information about fertility |
| 65. | Garvelink ^[65]^ | Study protocol on Garvelink, et al. ^[66]^ |
| 66. | Garvelink, et al. ^[66]^ | The study did not investigate the equality of the intervention effect between different social groups |
| 67. | Gattellari and Ward ^[67]^ | The study did not investigate the equality of the intervention effect between different social groups |
| 68. | Gebhard, et al. ^[68]^ | The study did not investigate the equality of the intervention effect between different social groups |
| 69. | Genz, et al. ^[69]^ | The study did not investigate the equality of the intervention effect between different social groups |
| 70. | Genz, et al. ^[70]^ | Wrong patient population |
| 71. | Gingras-Charl, et al. ^[71]^ | Wrong intervention (multicomponent intervention) |
| 72. | Goel, et al. ^[72]^ | The study did not investigate the equality of the intervention effect between different social groups |
| 73. | Gokce, et al. ^[73]^ | The study did not investigate the equality of the intervention effect between different social groups |
| 74. | Gong, et al. ^[74]^ | The study did not investigate the equality of the intervention effect between different social groups |
| 75. | Guillen, et al. ^[75]^ | The study did not investigate the equality of the intervention effect between different social groups |
| 76. | Gummersbach, et al. ^[76]^ | Study protocol on Gummersbach, et al. ^[77]^ |
| 77. | Gummersbach, et al. ^[77]^ | The study did not investigate the equality of the intervention effect between different social groups |
| 78. | Guo, et al. ^[78]^ | The study did not investigate the equality of the intervention effect between different social groups |
| 79. | Haakenson, et al. ^[79]^ | The study did not investigate the equality of the intervention effect between different social groups |
| 80. | Hamdiui, et al. ^[80]^ | Wrong patient population (focus on a specific disadvantaged group (Turkish- and Moroccan-Dutch women) |
| 81. | Heller, et al. ^[81]^ | The study did not investigate the equality of the intervention effect between different social groups |
| 82. | Hess, et al. ^[82]^ | Same study as Rising, et al. ^[83]^ (included in data synthesis) |
| 83. | Hoffman, et al. ^[84]^ | Wrong patient population (focus on a specific disadvantaged group (African American) |
| 84. | Hong, et al. ^[85]^ | No benefit-harm information on facial plastic surgery (only risks are reported) |
| 85. | Hu, et al. ^[86]^ | The study did not investigate the equality of the intervention effect between different social groups |
| 86. | Hua, et al. ^[87]^ | The study did not investigate the equality of the intervention effect between different social groups |
| 87. | Humphris and Field ^[88]^ | The study did not investigate the equality of the intervention effect between different social groups |
| 88. | Humphris, et al. ^[89]^ | The study did not investigate the equality of the intervention effect between different social groups |
| 89. | Humphris, et al. ^[90]^ | Wrong patient population |
| 90. | Hwang ^[91]^ | The study did not investigate the equality of the intervention effect between different social groups |
| 91. | Jack, et al. ^[92]^ | Wrong intervention (multicomponent intervention) |
| 92. | Jessop, et al. ^[93]^ | Wrong intervention (multicomponent intervention) |
| 93. | Jimenez, et al. ^[94]^ | Wrong outcome for subgroup analysis |
| 94. | Johnson, et al. ^[95]^ | The study did not investigate the equality of the intervention effect between different social groups |
| 95. | Kakkilaya, et al. ^[96]^ | The study did not investigate the equality of the intervention effect between different social groups |
| 96. | Kasper, et al. ^[97]^ | The study did not investigate the equality of the intervention effect between different social groups |
| 97. | Katz, et al. ^[98]^ | Wrong outcome (focus on uptake) |
| 98. | Kellar, et al. ^[99]^ | The study did not investigate the equality of the intervention effect between different social groups |
| 99. | Kennedy, et al. ^[100]^ | The study did not investigate the equality of the intervention effect between different social groups |
| 100. | Kim ^[101]^ | Study protocol without publication of results; just a further study protocol was found |
| 101. | Kim and Gong ^[102]^ | Two different evidence-based formats were tested (written vs. video) |
| 102. | Klifto, et al. ^[103]^ | The study did not investigate the equality of the intervention effect between different social groups |
| 103. | Knops, et al. ^[104]^ | The study did not investigate the equality of the intervention effect between different social groups |
| 104. | Krishnamurti, et al. ^[105]^ | The study did not investigate the equality of the intervention effect between different social groups |
| 105. | Krist, et al. ^[106]^ | The study did not investigate the equality of the intervention effect between different social groups |
| 106. | Kuppermann, et al. ^[107]^ | The study did not investigate the equality of the intervention effect between different social groups |
| 107. | Kuppermann, et al. ^[108]^ | Wrong intervention |
| 108. | Lai and Chan ^[109]^ | Wrong intervention (video vs. verbal information) |
| 109. | Lam, et al. ^[110]^ | The study did not investigate the equality of the intervention effect between different social groups |
| 110. | Lattuca, et al. ^[111]^ | Wrong intervention (written vs. video information) |
| 111. | Laupacis, et al. ^[112]^ | The study did not investigate the equality of the intervention effect between different social groups |
| 112. | LeCompte, et al. ^[113]^ | Wrong publication format (conference abstract, no further publication) |
| 113. | Legare, et al. ^[114]^ | The study did not investigate the equality of the intervention effect between different social groups |
| 114. | Leung, et al. ^[115]^ | The study did not investigate the equality of the intervention effect between different social groups |
| 115. | Lo, et al. ^[116]^ | Wrong publication (only statistical analysis plan, no further publication) |
| 116. | Lopez-Olivo, et al. ^[117]^ | Wrong intervention (booklet and video contain same information) |
| 117. | Lowe, et al. ^[118]^ | The study did not investigate the equality of the intervention effect between different social groups |
| 118. | Macy, et al. ^[119]^ | Wrong intervention |
| 119. | Madden, et al. ^[120]^ | The study did not investigate the equality of the intervention effect between different social groups |
| 120. | Maggs, et al. ^[121]^ | The study did not investigate the equality of the intervention effect between different social groups |
| 121. | Makdessian, et al. ^[122]^ | Wrong intervention (no benefit-harm information on facial plastic surgery (authors limited the study to only one aspect of informed consent requirements)) |
| 122. | Mann, et al. ^[123]^ | Wrong intervention |
| 123. | Man-Son-Hing, et al. ^[124]^ | Wrong study design (subgroup analysis on both groups intervention and control) |
| 124. | Mansoor and Dowse ^[125]^ | The study did not investigate the equality of the intervention effect between different social groups |
| 125. | Marteau, et al. ^[126]^ | Wrong outcome (No effect analysis related to vulnerability and target outcome) |
| 126. | Mathieu, et al. ^[127]^ | The study did not investigate the equality of the intervention effect between different social groups |
| 127. | Mathieu, et al. ^[128]^ | The study did not investigate the equality of the intervention effect between different social groups |
| 128. | McCarthy, et al. ^[129]^ | The study did not investigate the equality of the intervention effect between different social groups |
| 129. | Merchant, et al. ^[130]^ | The study did not investigate the equality of the intervention effect between different social groups |
| 130. | Miller, et al. ^[131]^ | Wrong outcomes |
| 131. | Miller, et al. ^[132]^ | Wrong outcomes (increase of screening uptake) |
| 132. | Montgomery, et al. ^[133]^ | The study did not investigate the equality of the intervention effect between different social groups |
| 133. | Morr, et al. ^[134]^ | Wrong intervention (pamphlet to increase awareness about peripheral arterial disease) |
| 134. | Movaseghi Ardekani, et al. ^[135]^ | Wrong intervention (education on oral health, no decision situation) |
| 135. | Myers, et al. ^[136]^ | The study did not investigate the equality of the intervention effect between different social groups |
| 136. | Nagle, et al. ^[137]^ | The study did not investigate the equality of the intervention effect between different social groups |
| 137. | Nassar, et al. ^[138]^ | The study did not investigate the equality of the intervention effect between different social groups |
| 138. | Nathan, et al. ^[139]^ | Wrong study design (systematic review) |
| 139. | Navas, et al. ^[140]^ | The study did not investigate the equality of the intervention effect between different social groups |
| 140. | O'Cathain, et al. ^[141]^ | The study did not investigate the equality of the intervention effect between different social groups |
| 141. | O'Connor, et al. ^[142]^ | The study did not investigate the equality of the intervention effect between different social groups |
| 142. | O'Donoghue, et al. ^[143]^ | Wrong intervention (intervention comparison) |
| 143. | Ohman, et al. ^[144]^ | The study did not investigate the equality of the intervention effect between different social groups |
| 144. | Ota, et al. ^[145]^ | Wrong intervention (in intervention 1 was no control; in intervention 2 there was a control, but the intervention was not a leaflet) |
| 145. | Paris, et al. ^[146]^ | Decision situation at meta level rather than concrete |
| 146. | Partin, et al. ^[147]^ | The study did not investigate the equality of the intervention effect between different social groups |
| 147. | Patzer, et al. ^[148]^ | Study protocol on Patzer, et al. ^[149]^ (study included in data synthesis) |
| 148. | Peate and Sandhu ^[150]^ | Study protocol (ACTRN12620001032943) without publication of results; just a further study protocol was found |
| 149. | Peele, et al. ^[151]^ | The study did not investigate the equality of the intervention effect between different social groups |
| 150. | Peipert, et al. ^[152]^ | Wrong intervention (multicomponent intervention) |
| 151. | Pernod, et al. ^[153]^ | No benefit-harm information on the use of anticoagulants; the information used is not intended to support a decision for or against the use of anticoagulants, only how to use them correctly |
| 152. | Piredda, et al. ^[154]^ | The study did not investigate the equality of the intervention effect between different social groups |
| 153. | Piredda, et al. ^[155]^ | Same study as Piredda, et al. ^[154]^ |
| 154. | Politi, et al. ^[156]^ | The study did not investigate the equality of the intervention effect between different social groups |
| 155. | Pot, et al. ^[157]^ | Wrong intervention (multicomponent) |
| 156. | Poureslami, et al. ^[158]^ | Wrong comparator |
| 157. | Protz, et al. ^[159]^ | The study did not investigate the equality of the intervention effect between different social groups |
| 158. | Prunty, et al. ^[160]^ | The study did not investigate the equality of the intervention effect between different social groups |
| 159. | Raymer, et al. ^[161]^ | Wrong intervention (multicomponent intervention) |
| 160. | Raynes-Greenow, et al. ^[162]^ | The study did not investigate the equality of the intervention effect between different social groups |
| 161. | Reder and Kolip ^[163]^ | The study did not investigate the equality of the intervention effect between different social groups |
| 162. | Rimer, et al. ^[164]^ | The study did not investigate the equality of the intervention effect between different social groups |
| 163. | Roberts, et al. ^[165]^ | The study did not investigate the equality of the intervention effect between different social groups |
| 164. | Roberts, et al. ^[166]^ | The information is not intended to support a decision (e.g. regarding hypertension treatment), but is general information about hypertension (missing benefit-harm presentation (non-evidence-based health information)) |
| 165. | Rodrigue, et al. ^[167]^ | Multicomponent intervention; no information on the content of living-donor kidney transplant (LDKT) information and education (whether benefits and harms are reported) |
| 166. | Roland and Dixon ^[168]^ | Wrong study design (subgroup analysis on both groups intervention and control) |
| 167. | Rostom, et al. ^[169]^ | Wrong intervention (format comparison) |
| 168. | Roter, et al. ^[170]^ | Wrong intervention (multicomponent intervention) |
| 169. | Rothwell, et al. ^[171]^ | The study did not investigate the equality of the intervention effect between different social groups |
| 170. | Ruffin IV, et al. ^[172]^ | Wrong outcome (Equity analysis only in relation to the secondary outcome "preferred method for colorectal cancer screening" (primary outcome: colorectal cancer screening received yes/no)) |
| 171. | Ruparel, et al. ^[173]^ | The study did not investigate the equality of the intervention effect between different social groups |
| 172. | Ruzek, et al. ^[174]^ | Wrong patient population (focus on African Americans with limited literacy) |
| 173. | Sajeev, et al. ^[175]^ | Wrong study design (pilot study, no RCT on DAs) |
| 174. | Sajeev, et al. ^[176]^ | Conference paper on Sajeev, et al. ^[175]^ |
| 175. | Sauvé, et al. ^[177]^ | The information is not intended to prepare a decision (e.g. regarding pre-eclampsia treatment), but is general information about pre-eclampsia (missing benefit-harm presentation (non-evidence-based health information)) |
| 176. | Saver, et al. ^[178]^ | The study did not investigate the equality of the intervention effect between different social groups |
| 177. | Schonberg, et al. ^[179]^ | Wrong outcome (uptake not outcome of interest) |
| 178. | Schroy III, et al. ^[180]^ | Equity analysis not for primary outcome knowledge |
| 179. | Schwalm, et al. ^[181]^ | The study did not investigate the equality of the intervention effect between different social groups |
| 180. | Schwartz, et al. ^[182]^ | The study did not investigate the equality of the intervention effect between different social groups |
| 181. | Schwartz, et al. ^[183]^ | The study did not investigate the equality of the intervention effect between different social groups |
| 182. | Selea, et al. ^[184]^ | The study did not investigate the equality of the intervention effect between different social groups |
| 183. | Shorten, et al. ^[185]^ | The study did not investigate the equality of the intervention effect between different social groups |
| 184. | Shue, et al. ^[186]^ | Wrong comparator |
| 185. | Simula, et al. ^[187]^ | The study did not investigate the equality of the intervention effect between different social groups |
| 186. | Singh, et al. ^[188]^ | Study protocol on Singh, et al. ^[189]^ (included) |
| 187. | Skjøth, et al. ^[190]^ | The study did not investigate the equality of the intervention effect between different social groups |
| 188. | Smith, et al. ^[191]^ | The study did not investigate the equality of the intervention effect between different social groups |
| 189. | Smith, et al. ^[192]^ | The study did not investigate the equality of the intervention effect between different social groups |
| 190. | Snyder-Ramos, et al. ^[193]^ | Two different formats with the same content were tested (written vs. video) |
| 191. | Staley, et al. ^[194]^ | Wrong outcomes; wrong intervention |
| 192. | Steckelberg, et al. ^[195]^ | The study did not investigate the equality of the intervention effect between different social groups |
| 193. | Steckelberg, et al. ^[196]^ | The study did not investigate the equality of the intervention effect between different social groups |
| 194. | Stephens, et al. ^[197]^ | Wrong study design (structural equation modeling) |
| 195. | Stern, et al. ^[198]^ | The study did not investigate the equality of the intervention effect between different social groups |
| 196. | Stewart, et al. ^[199]^ | The study did not investigate the equality of the intervention effect between different social groups |
| 197. | Stortz, et al. ^[200]^ | The study did not investigate the equality of the intervention effect between different social groups |
| 198. | Street Jr, et al. ^[201]^ | No equity analysis on knowledge, they study reports that perceived involvement correlates with age and education, but not in relation to the intervention |
| 199. | Sustersic, et al. ^[202]^ | Wrong intervention (information is not intended to support a decision (e.g. regarding gastroenteritis or tonsillitis treatment), but is general information about the respective diseases (missing benefit-harm presentation (non-evidence-based health information)) |
| 200. | Tomko, et al. ^[203]^ | The study did not investigate the equality of the intervention effect between different social groups |
| 201. | Tsai, et al. ^[204]^ | Wrong intervention |
| 202. | Tschautscher, et al. ^[205]^ | The study did not investigate the equality of the intervention effect between different social groups |
| 203. | Tucholka, et al. ^[206]^ | The study did not investigate the equality of the intervention effect between different social groups |
| 204. | Vanegas, et al. ^[207]^ | No publication on study data |
| 205. | Vilella, et al. ^[208]^ | No publication on study data |
| 206. | Vina, et al. ^[209]^ | Wrong patient population |
| 207. | Vodermaier, et al. ^[210]^ | The study did not investigate the equality of the intervention effect between different social groups |
| 208. | Volk, et al. ^[211]^ | Wrong patient population |
| 209. | Volk, et al. ^[212]^ | Wrong population (focus on patient with low health literacy) |
| 210. | Volk, et al. ^[213]^ | The study did not investigate the equality of the intervention effect between different social groups |
| 211. | Vuorma, et al. ^[214]^ | The study did not investigate the equality of the intervention effect between different social groups |
| 212. | Wakefield, et al. ^[215]^ | The study did not investigate the equality of the intervention effect between different social groups |
| 213. | Waterman, et al. ^[216]^ | Wrong intervention |
| 214. | Waterman, et al. ^[217]^ | Multicomponent intervention |
| 215. | Watkins, et al. ^[218]^ | No benefit-harm presentation (non-evidence-based health information) |
| 216. | Watson, et al. ^[219]^ | The study did not investigate the equality of the intervention effect between different social groups |
| 217. | Watts, et al. ^[220]^ | Wrong comparator |
| 218. | Weng, et al. ^[221]^ | Multicomponent intervention |
| 219. | Westerman, et al. ^[222]^ | No publication on study data |
| 220. | Weymiller, et al. ^[223]^ | The study did not investigate the equality of the intervention effect between different social groups |
| 221. | Wilhelm, et al. ^[224]^ | Wrong intervention (interactive DVD in addition to direct conversation) |
| 222. | Wilkens, et al. ^[225]^ | The study did not investigate the equality of the intervention effect between different social groups |
| 223. | Wilkie, et al. ^[226]^ | The study did not investigate the equality of the intervention effect between different social groups |
| 224. | Wolf, et al. ^[227]^ | The study did not investigate the equality of the intervention effect between different social groups (only summarised for various vulnerability dimensions and in relation to the overall sample) |
| 225. | Yee, et al. ^[228]^ | Wrong intervention (multicomponent intervention) |
| 226. | You, et al. ^[229]^ | Wrong intervention (multicomponent intervention) |

**Study protocols excluded with reasons**

|  | **Study protocols** | **Exclusion reason** |
| --- | --- | --- |
| 1. | DRKS00024850 | Study protocol on a trial (Debbeler, et al. ^[230]^, with comparison of two evidence-based formats (text vs. graphical) (exclusion: wrong comparison) |
| 2. | DRKS00029622: Health information management at the point of care | Study protocol without published data |
| 3. | Nct ^[232]^ | Study protocol for a Cochrane Review |
| 4. | NCT05800483 Nct ^[233]^: Improving Decision-Making for Low Health Literate Prostate CA Patients | Study protocol on a trial (Diefenbach, et al. ^[234]^), with comparison of two decision interventions (exclusion: wrong intervention type) |
| 5. | ACTRN12621000515897 Peate ^[235]^: Evaluating fertility decision aids for younger women with breast cancer. | Study protocol without published data |
| 6. | ACTRN12621000235808 Isautier ^[236]^ | Supporting Adults with Chronic Kidney Disease to engage in shared decision making successfully (SUCCESS): A Pragmatic randomised controlled trial of the SUCCESS intervention.  Wrong intervention (multicomponent) |

143. O'Donoghue AC, Sullivan HW, Aikin KJ, et al. Presenting efficacy information in direct-to-consumer prescription drug advertisements. *Patient Educ Couns*;95(2):271-80.

144. Ohman SG, Bjorklund U, Marsk A. Does an informational film increase women's possibility to make an informed choice about second trimester ultrasound? *Prenat Diagn*;32(9):833-9.

213. Volk RJ, Spann SJ, Cass AR, et al. Patient education for informed decision making about prostate cancer screening: A randomized controlled trial with 1-year follow-up. *Ann Fam Med*;1(1):22-8.

<https://www.ncbi.nlm.nih.gov/pmc/articles/PMC4043849/pdf/nihms574826.pdf> (accessed 2014-1-1).

229. You WB, Wolf MS, Bailey SC, et al. Improving patient understanding of preeclampsia: A randomized controlled trial. *Am J Obstet Gynecol*;206(5):431.e1-5.
