## Supplement 2: Tables S1-8 for "Can health information and decision aids decrease inequity in health care? A systematic review on the equality of their effectiveness"

**Table S1: Outcomes assessed in included studies (N=12).**

|  | **Knowledge** | **Decisional conflict** | **Intention/**  **Uptake** | **SDM** | **Informed decision/**  **decision concord.** | **Decision regret** | **Attitude** |
| --- | --- | --- | --- | --- | --- | --- | --- |
| Durand et al. 2021 | X |  | X | X | X | X |  |
| Gordon et al. 2017 | X |  |  |  |  |  |  |
| Healton et al. 1999 | X |  | X |  |  |  | X |
| Hewison et al. 2001 | X |  |  |  |  |  |  |
| Hunter et al. 2005 | X | X |  |  |  |  |  |
| Patzer et al. 2018 | X |  |  |  |  |  |  |
| Rising et al. 2018 | X | X |  | X |  |  |  |
| Singh et al. 2019 |  | X |  |  | X |  |  |
| Skains et al. 2019 | X | X |  | X |  |  |  |
| Thomas et al. 2013 | X | X | X |  |  |  |  |
| Trevena et al. 2008 | X |  |  |  |  |  |  |
| Williams et al. 2013 | X | X |  |  |  |  |  |
|  | **11** | **6** | **3** | **3** | **2** | **1** | **1** |

**Table S2: Inequity generating factors by decision-relevant outcomes in included studies (N=12).**

|  | Education | Ethnicity/  language | Age | Gender | HL | Numeracy | Income | SES | Graphical literacy | Employment status | Insurance status | Overall |
| --- | --- | --- | --- | --- | --- | --- | --- | --- | --- | --- | --- | --- |
| Knowledge | 5 | 7 | 4 | 4 | 4 | 3 | 3 | 2 |  | 1 | 1 | 34 |
| SDM | 2 | 2 | 1 | 2 | 2 | 2 | 2 | 2 |  |  | 1 | 16 |
| Decisional conflict | 3 | 5 | 1 | 2 | 3 | 3 | 3 | 1 | 1 |  | 1 | 23 |
| Informed decision/  decision concordance | 2 | 1 |  |  | 1 | 1 | 1 | 1 | 1 |  |  | 8 |
| Attitude | 1 | 1 | 1 |  |  |  |  |  |  |  |  | 3 |
| Intention/  uptake | 2 | 2 | 1 |  |  |  |  |  |  |  |  | 4 |
| Decision regret | 1 |  |  |  |  |  |  |  |  |  |  | 1 |

**Table S3. Study outcomes by inequality factor age.**

| **Outcome** | **Groups** | **Intervention condition** | | | **Control condition** | | | **Difference-in-differences** | **Post IV vs control**  **per group** | | **Post IA IV**  **x group** |
| --- | --- | --- | --- | --- | --- | --- | --- | --- | --- | --- | --- |
|  |  | **Baseline M(SD)** | **Post M(SD)** | **p** | **Baseline M(SD)** | **Post M(SD)** | **p** | **M (95%CI)** | **p** | **OR/M**  **(95%CI)** | **p** |
| **Knowledge (n=4)** | |  |  |  |  |  |  |  |  |  |  |
| Gordon 2017 | <= 40 years |  | 22.49 (1.07) |  |  | 15.49 (0.92) |  |  | <.001 |  |  |
|  | 41-50 years |  | 22.02 (1.15) |  |  | 14.42 (0.81) |  |  | <.001 |  |  |
|  | 51-60 years |  | 19.41 (0.91) |  |  | 13.58 (0.75) |  |  | <.001 |  |  |
|  | > 60 years |  | 20.04 (1.30) |  |  | 12.44 (0.78) |  |  | <.001 |  |  |
| Healton 1999 | 14-19 years |  |  |  |  |  |  |  |  |  | <.010 |
|  | >= 20 years |  |  |  |  |  |  |  |  |  |  |
| Hewison 2001 | Younger age groups |  |  |  |  |  |  |  |  |  | >.050 |
|  | Older age groups |  |  |  |  |  |  |  |  |  |  |
| Rising 2018 | <= 50 years |  |  |  |  |  |  |  |  | M=9.74  (6.31,13.17) | .236 |
|  | > 50 years |  |  |  |  |  |  |  |  | M=6.77  (3.18,10.36) |  |
| **Decisional conflict (n=1)** | |  |  |  |  |  |  |  |  |  |  |
| Rising 2018 | <= 50 years |  |  |  |  |  |  |  |  | M=-3.79  (-6.81,- 0.76) | .473 |
|  | > 50 years |  |  |  |  |  |  |  |  | M=-2.18  (-4.76,0.41) |  |
| **SDM (n=1)** |  |  |  |  |  |  |  |  |  |  |  |
| Rising 2018 | <= 50 years |  |  |  |  |  |  |  |  | M=10.31  (8.46,12.17) | .936 |
|  | > 50 years |  |  |  |  |  |  |  |  | M=10.23  (8.64,11.82) |  |

Note. IA = interaction, ii = ineffective intervention overall, IV = intervention, M = mean, OR = Odds ratio.

**Table S4. Study outcomes by inequality factor education.**

| **Outcome** | **Groups** | **Intervention condition** | | | **Control condition** | | | **Difference-in-differences** | **Post IV vs control**  **per group** | | **Post IA IV**  **x group** |
| --- | --- | --- | --- | --- | --- | --- | --- | --- | --- | --- | --- |
|  |  | **Baseline M(SD)** | **Post M(SD)** | **p** | **Baseline M(SD)** | **Post M(SD)** | **p** | **M (95%CI)** | **p** | **OR/M**  **(95%CI)** | **p** |
| **Knowledge (n=5)** |  |  |  |  |  |  |  |  |  |  |  |
| Gordon 2017 | <HS or less |  | 19.90 (0.63) |  |  | 12.65 (0.43) |  |  | <.001 |  |  |
|  | Some college or higher |  | 22.81 (1.08) |  |  | 17.20 (0.79) |  |  | <.001 |  |  |
| Healton 1999 | Less than HS |  |  |  |  |  |  |  |  |  | >.050 |
|  | HS |  |  |  |  |  |  |  |  |  |  |
|  | More than HS |  |  |  |  |  |  |  |  |  |  |
| Rising 2018 | <= HS |  |  |  |  |  |  |  |  | M=5.17  (0.61,9.73) | .059 |
|  | > HS |  |  |  |  |  |  |  |  | M=10.31  (7.35,13.27) |  |
| Skains 2019 | <or equal to HS/GED |  |  |  |  |  |  |  |  | OR=6.49  (0.55,13.53) | .619 |
|  | > HS/ GED |  |  |  |  |  |  |  |  | OR=9.46  (6.85,12.07) |  |
| Trevena 2008 | 16 years of school education or less |  | 50.0% |  |  | 17.8% |  |  |  |  | >.050 |
|  | Secondary school beyond age 16 |  | 31.3% |  |  | 19.4% |  |  |  |  |  |
|  | Tertiary education |  | 79.4% |  |  | 32.1% |  |  |  |  |  |
| **Behavioural Intention (n=1)** |  |  |  |  |  |  |  |  |  |  |  |
| Healton 1999 | Less than HS |  |  |  |  |  |  |  |  |  | >.050 |
|  | HS |  |  |  |  |  |  |  |  |  |  |
|  | More than HS |  |  |  |  |  |  |  |  |  |  |
| **Informed choice (n=1)** | |  |  |  |  |  |  |  |  |  |  |
| Singh 2019 | HS or less |  |  |  |  |  |  |  | .992 |  |  |
|  | Greater than HS |  |  |  |  |  |  |  | .103 |  |  |
| **Decisional conflict (n=2)** | |  |  |  |  |  |  |  |  |  |  |
| Singh 2019 | HS or less |  | 18.54 (3.43) |  |  | 11.40 (3.19) |  |  | .020 | M=9.20  (1.20,17.50) |  |
|  | Greater than HS |  | 23.60 (3.34) |  |  | 13.50 (2.61) |  |  | <.001 | M=12.70  (7.00,18.5) |  |
| Skains 2019 | <or equal to HS/GED |  |  |  |  |  |  |  |  | OR=-6.16  (-12.18,-0.14) | .375 |
|  | > HS/GED |  |  |  |  |  |  |  |  | OR=-4.02  (-6.26,-1.78) |  |
| **Decision regret (n=1)** | |  |  |  |  |  |  |  |  |  |  |
| Durand 2021 | <HS diploma or equivalent |  |  |  |  |  |  |  |  | M=50.56 (17.03,84.08) | .003 |
|  | > HS diploma or equivalent |  |  |  |  |  |  |  |  |  |  |
| **Attitudes (n=1)** |  |  |  |  |  |  |  |  |  |  |  |
| Healton 1999 | Less than HS |  |  |  |  |  |  |  |  |  | <.050 * |
|  | HS |  |  |  |  |  |  |  |  |  |  |
|  | More than HS |  |  |  |  |  |  |  |  |  |  |
| **SDM (n=2)** |  |  |  |  |  |  |  |  |  |  |  |
| Rising 2018 | <HS |  |  |  |  |  |  |  |  | M=10.87  (8.85,12.90) | .552 |
|  | > HS |  |  |  |  |  |  |  |  | M=10.04  (8.53,11.56) |  |
| Skains 2019 | <or equal to HS/GED |  | 6.49 (0.55,13.53) |  |  |  |  |  |  | OR=12.55  (9.70,15.40) | .762 |
|  | > HS/GED |  | 9.46 (6.85,12.07) |  |  |  |  |  |  | OR=11.81 (10.30,13.33) |  |

Note. GED = General Education Development, HS = High School, IA = interaction, ii = ineffective intervention overall, IV = intervention, M = mean, OR = Odds ratio, * this study does not report any adjustment of p-values despite 42 tests for possible covariates.

**Table S5. Study outcomes by inequality factors race, ethnicity and language preference.**

| **Outcome** | **Groups** | **Intervention condition** | | | **Control condition** | | | **Difference-in-differences** | **Post IV vs control**  **per group** | | **Post IA IV**  **x group** |
| --- | --- | --- | --- | --- | --- | --- | --- | --- | --- | --- | --- |
|  |  | **Baseline M(SD)** | **Post M(SD)** | **p** | **Baseline M(SD)** | **Post M(SD)** | **p** | **M (95%CI)** | **p** | **OR/M**  **(95%CI)** | **p** |
| **Race and ethnicity** | |  |  |  |  |  |  |  |  |  |  |
| **Knowledge (n=7)** | |  |  |  |  |  |  |  |  |  |  |
| Gordon 2017 | Non-hispanic W |  | 23.36 (0.94) |  |  | 15.88 (0.81) |  |  | <.001 |  |  |
|  | Non-hispanic Black |  | 19.57 (0.74) |  |  | 13.15 (0.47) |  |  | <.001 |  |  |
|  | Other |  | 18.97 (1.46) |  |  | 12.30 (1.33) |  |  | .002 |  |  |
| Healton 1999 | Non-Latina W |  |  |  |  |  |  |  |  |  | <.010 |
|  | African American |  |  |  |  |  |  |  |  |  |  |
|  | Puerto Rican |  |  |  |  |  |  |  |  |  |  |
|  | Other Latina |  |  |  |  |  |  |  |  |  |  |
|  | Other |  |  |  |  |  |  |  |  |  |  |
| Patzer 2018 | African American | 4.68 (2.03) | 5.81 (2.02) |  | 4.60 (2.16) | 5.10 (1.70) |  | 0.63  (0.17,1.09 ) | .007 |  |  |
|  | Hispanic W | 4.64 (2.14) | 6.16 (1.82) |  | 4.55 (2.23) | 4.45 (2.18) |  | 1.61  (0.37,2.85) | .010 |  |  |
|  | Non-hispanic W | 5.69 (2.05) | 6.57 (1.77) |  | 5.94 (1.89) | 6.21 (1.70) |  | 0.60  (0.09,1.19) | .046 |  |  |
| Rising 2018 | Caucasian W |  |  |  |  |  |  |  |  | M=10.95  (7.76,14.14) | .018 |
|  | Non-W |  |  |  |  |  |  |  |  | M=4.79  (0.89,8.70) |  |
| Skains 2019 | W |  |  |  |  |  |  |  |  | OR=9.70  (6.91,12.48) | .774 |
|  | Non-W |  |  |  |  |  |  |  |  | OR=5.91  (0.84,10.97) |  |
| Thomas 2013 | W Patients | 8.8 (2.6) | 11.2 (1.3) | <.010 | 7.7 (4.2) | 9.5 (3.5) | <.010 |  | ii |  |  |
|  | Black Patients | 7.7 (2.9) | 10.1 (2.9) | <.010 | 6.7 (3.5) | 9.9 (1.3) | <.010 |  | ii |  |  |
| Williams 2013 | African Americans |  |  |  |  |  |  |  |  |  | n.s. |
|  | W |  |  |  |  |  |  |  |  |  |  |
| **Behavioural intentions [n=1]** | | | |  |  |  |  |  |  |  |  |
| Healton 1999 | Non-Latina W |  |  |  |  |  |  |  |  |  | > .050 |
|  | African American |  |  |  |  |  |  |  |  |  |  |
|  | Puerto Rican |  |  |  |  |  |  |  |  |  |  |
|  | Other Latina |  |  |  |  |  |  |  |  |  |  |
| **Informed choice (n=1)** | |  |  |  |  |  |  |  |  |  |  |
| Singh 2019 | Non-Hispanic Black |  |  |  |  |  |  |  | 0.216 |  |  |
|  | Hispanic/Latino |  |  |  |  |  |  |  | 0.612 |  |  |
|  | Non-Hispanic W |  |  |  |  |  |  |  | 0.003 |  |  |
|  | Asian/Other |  |  |  |  |  |  |  | 0.625 |  |  |
| **Decision making [n=1]** | |  |  |  |  |  |  |  |  |  |  |
| Thomas 2013 | W Patients |  | 79.2 |  |  | 84.6 |  |  | ii |  |  |
|  | Black Patients |  | 60.0 |  |  | 42.9 |  |  | ii |  |  |
| **Decisional conflict [n=5]** | |  |  |  |  |  |  |  |  |  |  |
| Skains 2019 | W |  |  |  |  |  |  |  |  | OR=-3.07  (-5.45,-0.68) | .050 |
|  | Non-W |  |  |  |  |  |  |  |  | OR=-8.14  (-12.33,-3.95) |  |
| Rising 2018 | W/Caucasian |  |  |  |  |  |  |  |  | M=-2.79  (-5.19,- 0.39) | .863 |
|  | NonW |  |  |  |  |  |  |  |  | M=-2.96  (-6.39,0.47) |  |
| Thomas 2013 | W Patients |  |  |  |  |  |  |  | ii |  |  |
|  | Black Patients |  |  |  |  |  |  |  | ii |  |  |
| Singh 2019 | Non-Hispanic Black |  | 25.52 (3.71) |  |  | 16.99 (3.18) |  |  | <.001 | M=11.8  (4.9,18,7) |  |
|  | Hispanic/Latino |  | 13.5 (4.29) |  |  | 6.76 (3.99) |  |  | .070 | M=8.8  (-0.9,18.6) |  |
|  | W, Non-Hispanic |  | 30.25 (8.6) |  |  | 12.29 (3.85) |  |  | .002 | M=19  (7.2,30.8) |  |
|  | Asian/Other |  | 19.17 (6.34) |  |  | 7.44 (4.61) |  |  | .050 | M=12.8  (-0.2,25.7) |  |
| Williams 2013 | African Americans |  |  |  |  |  |  |  | ii |  |  |
|  | W |  |  |  |  |  |  |  | ii |  |  |
| **SDM (n=2)** | |  |  |  |  |  |  |  |  |  |  |
| Rising 2018 | W/Caucasian |  |  |  |  |  |  |  |  | M=10.75  (9.28,12.22) | .316 |
|  | NonW |  |  |  |  |  |  |  |  | M=9.42  (7.27,11.57) |  |
| Skains 2019 | W |  |  |  |  |  |  |  |  | OR=12.27  (10.75,13.79) | .170 |
|  | Non-W |  |  |  |  |  |  |  |  | OR=9.98 (7.29,12.67) |  |
| **Language preference** | |  |  |  |  |  |  |  |  |  |  |
| **Knowledge (n=1)** | |  |  |  |  |  |  |  |  |  |  |
| Healton 1999 | Mono Spanish |  |  |  |  |  |  |  |  |  | <.010 |
|  | Bilingual |  |  |  |  |  |  |  |  |  |  |
|  | English |  |  |  |  |  |  |  |  |  |  |
| **Behavioural intention (n=1)** | | |  |  |  |  |  |  |  |  |  |
| Healton et al. 1999 | Mono Spanish |  |  |  |  |  |  |  |  |  | <.050 |
|  | Bilingual |  |  |  |  |  |  |  |  |  |  |
|  | English |  |  |  |  |  |  |  |  |  |  |
| **Decisional conflict (n=1)** | |  |  |  |  |  |  |  |  |  |  |
| Singh 2019 | Spanish |  | 19.77 (6.19) |  |  | 9.75 (6.18) |  |  | .120 |  |  |
|  | English |  | 22.15 (2.77) |  |  | 13.13 (2.13) |  |  | <.001 | M=11.9  (6.9,16.9) |  |
| **Informed choice (n=1)** | |  |  |  |  |  |  |  |  |  |  |
| Singh 2019 | Spanish |  |  |  |  |  |  |  | .898 |  |  |
|  | English |  |  |  |  |  |  |  | .059 |  |  |

Note. IA = interaction, ii = ineffective intervention overall, IV = intervention, M = mean, n.s. = not significant, OR = Odds ratio, W = Whites

**Table S6. Study outcomes by inequality factor gender.**

| **Outcomes** | **Groups** | **Intervention condition** | | | **Control condition** | | | **Difference-in-differences** | **Post IV vs control**  **per group** | | **Post IA IV**  **x group** |
| --- | --- | --- | --- | --- | --- | --- | --- | --- | --- | --- | --- |
|  |  | **Baseline M(SD)** | **Post M(SD)** | **p** | **Baseline M(SD)** | **Post M(SD)** | **p** | **M (95%CI)** | **p** | **OR/M**  **(95%CI)** | **p** |
| **Knowledge (n=4)** |  |  |  |  |  |  |  |  |  |  |  |
| Gordon 2017 | Female |  | 20.07 (0.85) |  |  | 14.10 (0.69) |  |  | <.001 |  |  |
|  | Male |  | 21.08 (0.73) |  |  | 13.85 (0.52 |  |  | <.001 |  |  |
| Hunter 2005 | Female | 15.51 (5.96) | 24.52 (7.45) |  | 15.99 (6.81) | 22.85 (7.22) |  |  |  |  |  |
|  | Male | 13.21 (6.10) | 22.44 (7.77) |  | 13.25 (6.88) | 20.51 (7.39) |  |  |  |  |  |
| Patzer 2018 | Female | 5.35 (1.93) | 6.14 (1.75) |  | 5.40 (2.16) | 5.57 (1.82) |  | 0.63 (0.12,1.14 ) | .020 |  |  |
|  | Male | 4.83 (2.21) | 6.09 (2.01) |  | 4.92 (2.15) | 5.43 (1.91) |  | 0.76 (0.32,1.19) | <.001 |  |  |
| Rising 2018 | Female |  |  |  |  |  |  |  |  | M=8.18  (4.95,11.40) | .928 |
|  | Male |  |  |  |  |  |  |  |  | M=8.48  (4.57,12.39) |  |
| **Decisional conflict (n=2)** | |  |  |  |  |  |  |  |  |  |  |
| Rising 2018 | Female |  |  |  |  |  |  |  |  | M=-3.47  (-6.01,- 0.94) | .439 |
|  | Male |  |  |  |  |  |  |  |  | M=-2.06  (-5.21,1.08) |  |
| Hunter 2005 | Female | 2.40 (0.58) | 1.73 (0.50) |  | 2.41 (0.56) | 1.85 (0.50) |  |  |  |  |  |
|  | Male | 2.35 (0.55) | 1.73 (0.45) |  | 2.48 (0.65) | 1.90 (0.50) |  |  |  |  |  |
| **SDM (n=1)** |  |  |  |  |  |  |  |  |  |  |  |
| Rising 2018 | Female |  |  |  |  |  |  |  |  | M=10.68  (9.13,12.22) | .420 |
|  | Male |  |  |  |  |  |  |  |  | M=9.67  (7.74,11.60) |  |

Note. IA = interaction, ii = ineffective intervention overall, IV = intervention, M = mean, OR = Odds ratio

**Table S7. Study outcomes by inequality factors health literacy, graphical literacy, and numeracy.**

| **Outcomes** | **Groups** | | **Intervention condition** | | | | **Control condition** | | | **Difference-in-differences** | | **Post IV vs control per group** | | **Post IA IV**  **x group** |
| --- | --- | --- | --- | --- | --- | --- | --- | --- | --- | --- | --- | --- | --- | --- |
|  |  | | **Baseline M(SD)** | **Post M(SD)** | | **p** | **Baseline M(SD)** | **Post M(SD)** | **p** | **M (95%CI)** | | **p** | **OR/M**  **(95%CI)** | **p** |
| **Health literacy** | | |  |  | |  |  |  |  |  | |  |  |  |
| **Knowledge (n=4)** | | |  |  | |  |  |  |  |  | |  |  |  |
| Patzer 2018 | Low  (0-1 points) | | 3.96 (2.02) | 5.06 (2.07) | |  | 4.20 (1.94) | 4.83 (1.77) |  | 0.47  (-0.36,1.31) | | .260 |  |  |
|  | Medium  (2-3 points) | | 4.98 (2.04) | 6.19 (1.86) | |  | 4.32 (2.09) | 4.91 (2.04) |  | 0.62  (0.04,1.19) | | .040 |  |  |
|  | High  (4-6 points) | | 5.53 (2.04) | 6.57 (1.66) | |  | 5.74 (2.05) | 5.97 (1.70) |  | 0.60  (0.09,1.19) | | <.001 |  |  |
| Gordon 2017 | Adequate | |  | 22.30 (0.73) | |  |  | 15.36 (0.55) |  |  | | <.001 |  |  |
|  | Moderate | |  | 19.70 (0.88) | |  |  | 12.04 (0.73) |  |  | | <.001 |  |  |
|  | Inadequate | |  | 17.76 (1.33) | |  |  | 12.21 (0.85) |  |  | | <.001 |  |  |
| Rising 2018 | Low (>3) | |  |  | |  |  |  |  |  | |  | M=6.67  (3.63,9.71) | .095 |
|  | Typical (3) | |  |  | |  |  |  |  |  | |  | M=11.02  (6.86,15.18) |  |
| Skains 2019 | Low | |  |  | |  |  |  |  |  | |  | OR=14.61 (7.36,21.85) | .063 |
|  | Typical | |  |  | |  |  |  |  |  | |  | OR=8.25  (5.60,10.90) |  |
| **Decisional conflict (n=3)** | |  | | |  | |  |  |  |  |  |  |  |  |
| Singh 2019 | Low | |  | 35 (8.26) | |  |  | 10.83 (7.25) |  |  | | .020 | M=19.5  (3.7,35.2) |  |
|  | High | |  | 20.9 (2.66) | |  |  | 12.85 (2.12) |  |  | | <.001 | M=11.2  (6.1,16.2) |  |
| Rising 2018 | Low (>3) | |  |  | |  |  |  |  |  | |  | M=-3.07  (-5.66,-0.47) | .829 |
|  | Typical (3) | |  |  | |  |  |  |  |  | |  | M=-2.47  (-5.38,0.45) |  |
| Skains 2019 | Low | |  |  | |  |  |  |  |  | |  | OR=-5.87  (-12.03,0.29) | .402 |
|  | Typical | |  |  | |  |  |  |  |  | |  | OR=-4.23  (-6.42,-2.04) |  |
| **Informed choice (n=1)** | |  | | |  | |  |  |  |  |  |  |  |  |
| Singh 2019 | Low | |  |  | |  |  |  |  |  | | .762 |  |  |
|  | High | |  |  | |  |  |  |  |  | | .091 |  |  |
| **SDM (n=2)** |  | |  |  | |  |  |  |  |  | |  |  |  |
| Rising 2018 | Low (>3) | |  |  | |  |  |  |  |  | |  | M=10.17 (8.67,11.66) | .812 |
|  | Typical (3) | |  |  | |  |  |  |  |  | |  | M=10.47 (8.43,12.52) |  |
| Skains 2019 | Low | |  |  | |  |  |  |  |  | |  | OR=11.78 (8.57,14.99) | .802 |
|  | Typical | |  |  | |  |  |  |  |  | |  | OR=11.85 (10.40,13.31) |  |
| **Graphical literacy** |  | |  |  | |  |  |  |  |  | |  |  |  |
| **Decisional conflict (n=1)** | |  | | |  | |  |  |  |  |  |  |  |  |
| Singh 2019 | Low | |  | 19.25 (2.71) | |  |  | 14.07 (2.26) |  |  | | <.001 | M=9.7 (4.1,15.2) |  |
|  | High | |  | 30.56 (5.99) | |  |  | 8.97 (4.39) |  |  | | .003 | M=14.6 (5.3,24.0) |  |
| **Informed choice (n=1)** | |  | | |  | |  |  |  |  |  |  |  |  |
| Singh 2019 | Low | |  |  | |  |  |  |  |  | | .684 |  |  |
|  | High | |  |  | |  |  |  |  |  | | .004 |  |  |
| **Numeracy** |  | |  |  | |  |  |  |  |  | |  |  |  |
| **Knowledge (n=3)** |  | |  |  | |  |  |  |  |  | |  |  |  |
| Rising 2018 | Low (>4): | |  |  | |  |  |  |  |  | |  | M=4.65 (0.83,8.47) | .025 |
|  | Typical (4): | |  |  | |  |  |  |  |  | |  | M=10.64 (7.45,13.84) |  |
| Skains 2019 | Low (<34) | |  |  | |  |  |  |  |  | |  | OR=6.50 (1.89,11.12) | .142 |
|  | Typical (≥34) | |  |  | |  |  |  |  |  | |  | OR=10.22 (7.36,13.08) |  |
| Patzer 2018 | Low (0-4 points) | | 3.96 (2.23) | 5.09 (2.09) | |  | 4.18 (2.13) | 4.85 (1.96) |  | 0.46  (-0.21,1.14) | | .180 |  |  |
|  | Medium (5-8) | | 5.23 (1.77) | 6.48 (1.59) | |  | 5.30 (2.09) | 5.60 (1.71) |  | 0.96  (0.45,1.47) | | <.001 |  |  |
|  | High (9-11) | | 6.00 (1.96) | 6.88 (1.51) | |  | 5.85 (1.95) | 6.05 (1.78) |  | 0.68  (0.12,1.25) | | .020 |  |  |
| **Decisional conflict (n=3)** | |  | | |  | |  |  |  |  |  |  |  |  |
| Rising 2018 | Low (>4): | |  |  | |  |  |  |  |  | |  | M=-2.47  (-4.95,0.02) | .454 |
|  | Typical (4): | |  |  | |  |  |  |  |  | |  | M=-3.97  (-7.17,-0.76) |  |
| Singh 2019 | Low | |  | 18.82 (4.71) | |  |  | 14.31 (4.33) |  |  | | .240 | M=6.8  (-4.7,18.4) |  |
|  | High | |  | 21.6 (3.72) | |  |  | 12.21 (2.99) |  |  | | <.001 | M=10.8 (4.8,16.7) |  |
| Skains 2019 | Low (<34) | |  |  | |  |  |  |  |  | |  | OR=-4.86  (-8.84,-0.88) | .741 |
|  | Typical (≥34) | |  |  | |  |  |  |  |  | |  | OR=-4.25  (-6.62,-1.88) |  |
| **Informed choice (n=1)** | |  | | |  | |  |  |  |  |  |  |  |  |
| Singh 2019 | Low | |  |  | |  |  |  |  |  | | .643 |  |  |
|  | High | |  |  | |  |  |  |  |  | | .006 |  |  |
| **SDM (n=2)** |  | |  |  | |  |  |  |  |  | |  |  |  |
| Rising 2018 | Low (>4): | |  |  | |  |  |  |  |  | |  | M=10.38 (8.82,11.94) | .885 |
|  | Typical (4): | |  |  | |  |  |  |  |  | |  | M=10.20 (8.24,12.16) |  |
| Skains 2019 | Low | |  |  | |  |  |  |  |  | |  | OR=11.72 (9.51,13.93) | .756 |
|  | Typical | |  |  | |  |  |  |  |  | |  | OR=11.51 (9.89,13.13) |  |

Note. IA = interaction, ii = ineffective intervention overall, IV = intervention, M = mean, OR = Odds ratio

**Table S8. Study outcomes by inequality factors of socioeconomic status.**

| **Outcomes** | **Groups** | **Intervention condition** | | **Control condition** | | | | **Difference-in-differences** | **Post IV vs control**  **per group** | | **Post IA IV**  **x group** |
| --- | --- | --- | --- | --- | --- | --- | --- | --- | --- | --- | --- |
|  |  | **Baseline M(SD)** | **Post M(SD)** | **p** | **Baseline M(SD)** | **Post M(SD)** | **p** | **M (95%CI)** | **p** | **OR/M**  **(95%CI)** | **p** |
| **Income** |  |  |  |  |  |  |  |  |  |  |  |
| **Knowledge (n=3)** | |  |  |  |  |  |  |  |  |  |  |
| Gordon 2017 | <$25,000 |  | 18.59 (0.77) |  |  | 12.78 (0.52) |  |  | <0.001 |  |  |
|  | $25,000 - $64,999 |  | 21.58 (1.00) |  |  | 13.73 (0.64) |  |  | <0.001 |  |  |
|  | >= $65,000 |  | 24.62 (0.83) |  |  | 17.67 (1.17) |  |  | <0.001 |  |  |
|  | Unknown |  | 21.54 (2.37) |  |  | 13.86 (1.99) |  |  | 0.02 |  |  |
| Rising 2018 | AI <$40,000 |  |  |  |  |  |  |  |  | M=6.76  (2.72,10.81) | .317 |
|  | AI >=$40,000 |  |  |  |  |  |  |  |  | M=9.46  (6.21,12.71) |  |
| Skains 2019 | AI <$40,000 |  |  |  |  |  |  |  |  | OR=9.53  (4.73,14.33) | .999 |
|  | AI >=$40,000 |  |  |  |  |  |  |  |  | OR=9.65  (6.85,12.45) |  |
| **Decisional conflict (n=3)** | |  |  |  |  |  |  |  |  |  |  |
| Rising 2018 | AI <$40,000 |  |  |  |  |  |  |  |  | M=-1.05  (-4.71,2.62) | .366 |
|  | AI >=$40,000 |  |  |  |  |  |  |  |  | M=-3.14  (-5.53,-0.76) |  |
| Singh 2019 | AI <$40,000 |  | 22.88 (3.38) |  |  | 13.13 (2.93) |  |  | <.001 | M=13.20  (6.60,19.90) |  |
|  | AI $40,000-$80,000 |  | 16.67 (5.24) |  |  | 16.41 (4.50) |  |  | .220 | M=5.80  (-3.7,15.3) |  |
|  | AI $80,000 or more |  | 32.06 (10.09) |  |  | 15.67 (6.32) |  |  | .047 | M=17.7  (0.2,35.1) |  |
| Skains 2019 | AI <$40,000 |  |  |  |  |  |  |  |  | OR=-6.18  (-10.13,-2.23) | .294 |
|  | AI >=$40,000 |  |  |  |  |  |  |  |  | OR=-3.55  (-6.00,-1.11) |  |
| **Informed choice (n=1)** | |  |  |  |  |  |  |  |  |  |  |
| Singh 2019 | AI <$40,000 |  |  |  |  |  |  |  | .317 |  |  |
|  | AI $40,000-$80,000 |  |  |  |  |  |  |  | .557 |  |  |
|  | AI $80,000 or more |  |  |  |  |  |  |  | .148 |  |  |
| **SDM (n=2)** |  |  |  |  |  |  |  |  |  |  |  |
| Rising 2018 | AI <$40,000 |  |  |  |  |  |  |  |  | M=10.06  (7.96,12.16) | .832 |
|  | AI >=$40,000 |  |  |  |  |  |  |  |  | M=10.25  (8.70,11.80) |  |
| Skains 2019 | <$40k |  |  |  |  |  |  |  |  | OR=12.90  (10.50,15.30) | .367 |
|  | ≥$40k |  |  |  |  |  |  |  |  | OR=11.35  (9.75,12.95) |  |
| **Socioeconomic status** | |  |  |  |  |  |  |  |  |  |  |
| **Knowledge (n=1)** | |  |  |  |  |  |  |  |  |  |  |
| Durand 2021 | Lower SES |  |  |  |  |  |  |  |  | M=0.36  (0.09,0.63) | .010 |
|  | Higher SES |  |  |  |  |  |  |  |  |  |  |
| **Decision concordance (n=1)** | |  |  |  |  |  |  |  |  |  |  |
| Durand 2021 | Lower SES |  |  |  |  |  |  |  |  |  | n.s. |
|  | Higher SES |  |  |  |  |  |  |  |  |  |  |
| **SDM (n=1)** | |  |  |  |  |  |  |  |  |  |  |
| Durand 2021 | Lower SES |  |  |  |  |  |  |  |  |  | n.s. |
|  | Higher SES |  |  |  |  |  |  |  |  |  |  |
| **Treatment choice (n=1)** | |  |  |  |  |  |  |  |  |  |  |
| Durand 2021 | Lower SES |  |  |  |  |  |  |  |  |  | n.s. |
|  | Higher SES |  |  |  |  |  |  |  |  |  |  |
| **Socioeconomically (dis-)advantaged** | | |  |  |  |  |  |  |  |  |  |
| **Knowledge (n=1)** | |  |  |  |  |  |  |  |  |  |  |
| Skains 2019 | SD |  |  |  |  |  |  |  |  | OR=6.65  (-1.10,14.39) | .298 |
|  | SA |  |  |  |  |  |  |  |  | OR=9.25  (6.63,11.87) |  |
| **Decisional conflict (n=1)** | |  |  |  |  |  |  |  |  |  |  |
| Skains 2019 | SD |  |  |  |  |  |  |  |  | OR=-6.41  (-12.19,-0.64) | .725 |
|  | SA |  |  |  |  |  |  |  |  | OR=-3.96  (-6.17,-1.75) |  |
| **SDM (n=1)** |  |  |  |  |  |  |  |  |  |  |  |
| Skains 2019 | SD |  |  |  |  |  |  |  |  | OR=10.57  (5.97,15.16) | .569 |
|  | SA |  |  |  |  |  |  |  |  | OR=11.86  (10.46,13.27) |  |
| **Employment status** |  |  |  |  |  |  |  |  |  |  |  |
| **Knowledge (n=1)** |  |  |  |  |  |  |  |  |  |  |  |
| Gordon 2017 | Not working |  | 19.23 (0.71) |  |  | 13.30 (0.45) |  |  | <.001 |  |  |
|  | Working |  | 23.54 (0.75) |  |  | 15.99 (0.92) |  |  | <.001 |  |  |
| **Health insurance status** | |  |  |  |  |  |  |  |  |  |  |
| **Knowledge (n=1)** |  |  |  |  |  |  |  |  |  |  |  |
| Rising 2018 | Uninsured |  |  |  |  |  |  |  |  | M=3.76 (-5.28,12.79) | .236 |
|  | Insured |  |  |  |  |  |  |  |  | M=8.72 (6.13,11.30) |  |
| **Decisional conflict (n=1)** | |  |  |  |  |  |  |  |  |  |  |
| Rising 2018 | Uninsured |  |  |  |  |  |  |  |  | M=-8.47 (-15.63,- 1.30) | .167 |
|  | Insured |  |  |  |  |  |  |  |  | M=-2.48 (-4.55,-0.41) |  |
| **SDM (n=1)** |  |  |  |  |  |  |  |  |  |  |  |
| Rising 2018 | Uninsured |  |  |  |  |  |  |  |  | M=10.67 (5.02,16.32) | .727 |
|  | Insured |  |  |  |  |  |  |  |  | M=10.36 (9.13,11.60) |  |

Note. AI = Annual income, IA = interaction, ii = ineffective intervention overall, IV = intervention, M = mean, OR = Odds ratio, SA = Socioeconomically advantaged, SD = Socioeconomically disadvantaged.
